# Facing the Omicron variant – How well do vaccines protect against mild and severe COVID-19? Third interim analysis of a living systematic review

**DOI:** 10.1101/2022.05.25.22275516

**Authors:** Wiebe Külper-Schiek, Vanessa Piechotta, Antonia Pilic, Madeleine Batke, Léa-Sophie Dreveton, Brogan Geurts, Judith Koch, Stefan Köppe, Marina Treskova, Sabine Vygen-Bonnet, Maria Waize, Ole Wichmann, Thomas Harder

**Author notes:** these authors have contributed equally to this work and share first authorship (Wiebe Külper-Schiek, Vanessa Piechotta).

## Abstract

**Background:** The SARS-CoV-2 Omicron variant is currently the dominant variant globally. This 3^rd^ interim analysis of a living systematic review summarizes evidence on COVID-19 vaccine effectiveness (VE) and duration of protection against Omicron.

**Methods:** We systematically searched the COVID-19 literature for controlled studies evaluating the effectiveness of COVID-19 vaccines approved in the European Union up to 14/01/2022, complemented by hand-searches of websites and metasearch engines up to 11/02/2022. We considered the following comparisons: full primary immunization vs. no vaccination; booster immunization vs. no vaccination; booster vs. primary immunization. VE against any confirmed SARS-CoV-2 infection, symptomatic, and severe COVID-19 (i.e. COVID-19-related hospitalization, ICU-admission, or death) was indicated providing estimate ranges. Meta-analysis was not performed due to high study heterogeneity. Risk of bias was assessed with ROBINS-I, certainty of evidence evaluated using GRADE.

**Results:** We identified 26 studies, including 430 to 2.2 million participants.

VE against any confirmed SARS-CoV-2 infection compared to no vaccination ranged between 0-62% after full primary immunization, and between 34-66% after a booster dose. VE-range for booster vs. primary immunization was 34-54.6%.

Against symptomatic COVID-19, VE ranged between 6-76% after full primary immunization, and between 19-73.9% after booster immunization, if compared to no vaccination. When comparing booster vs. primary immunization VE ranged between 56-69%.

VE against severe COVID-19 compared to no vaccination ranged between 3-84% after full primary immunization, and between 12-100% after a booster dose. One study compared booster vs. primary immunization (VE 100%, 95% CI 71.4-100).

VE was characterized by a moderate to strong decline within three to six months for SARS-CoV-2 infections and symptomatic COVID-19. Against severe COVID-19 protection remained robust at least for up to six months. Waning immunity was more profound after primary than booster immunization.

Risk of bias was moderate to critical across studies and outcomes. GRADE-certainty was very low for all outcomes.

**Author’s conclusions:** Under the Omicron variant, effectiveness of EU-licensed COVID-19 vaccines in preventing any SARS-CoV-2 infection or mild disease is low and only short-lasting after primary immunization, but can be improved by booster vaccination. VE against severe COVID-19 remains high and is long-lasting, especially after receiving the booster vaccination.

## 1 Introduction

The Omicron variant of SARS-CoV-2 (Phylogenetic Assignment of Named Global Outbreak (Pango) lineage designation B.1.1.529) was first detected in South-Africa in November 2021. Since then, the variant spread rapidly across countries and has largely replaced all other variants globally (>99,8% of all sequences submitted to the Global Initiative on Sharing All Influenza Data (GISAID) were Omicron in week 5 of 2022) (1). Evidence suggests that the Omicron variant has a growth rate advantage compared to the previously dominant Delta variant, leading to its overtake as the dominant variant globally, while the Delta variant now only represents 0.1% of collected samples (1).

High rates of asymptomatic infection and symptomatic COVID-19 disease among people previously infected with SARS-CoV-2 or fully vaccinated with a COVID-19 vaccine raise concerns that the currently available vaccines are less or no-longer effective against the Omicron variant. To summarize the existing evidence on the effectiveness and the duration of protection confered by COVID-19 vaccines licensed in the European Union (EU) with respect to the Omicron variant, and compare it to the Delta variant, we synthesized the evidence within an ongoing living systematic review (LSR), conducted by Robert Koch Institute (RKI) in collaboration with the National Immunisation Technical Advisory Groups (NITAGs) network coordinated by the European Centre for Disease Prevention and Control (ECDC) (2).

## 2 Methods

### Literature search

The LSR follows the Preferred Reporting Items for Systematic Review and Meta-Analysis (PRISMA) guideline (**supplement material 1, part 1**), and was registered in the *Prospective Register of Systematic Reviews* (PROSPERO registration ID: CRD42020208935; updated on: 09 March 2022). All amendments since initial registration are available online. The methods have been previously described in detail (2). In brief, we included studies of any design that had a comparison group and investigated vaccine effectiveness (VE) against SARS-CoV-2 infection of any severity of COVID-19 vaccines approved by the European Medicine Agency (EMA) in people ≥12 years of age (see **supplement material 1, part 2** for complete PICO question). For this 3^rd^ update of the LSR, we only considered studies that reported on outcomes that are due to the SARS-CoV-2 Omicron variant or that occurred during a dominant circulation of the Omicron variant. There were no restrictions with regard to publication language or status.

We searched the COVID-19 literature database constructed by the RKI library (see (2) and **supplement material 1, part 3** for description of the database and the search strategy) for studies published between 23 October 2021 and 14 January 2022. We added literature identified by hand-search of websites and metasearch engines indicated in **supplement material 1, part 3** up to 11 February 2022. Potentially relevant publications were screened at title/abstract and full-text level by pairs of independent investigators (AP, SK, MB, LSD, VP and/or WKS). Data from included studies (see PROSPERO registration for details) was extracted in duplicate and summarized in tables (AP, MB, VP, WKS). We considered VE data on the following three comparisons by vaccine type (mRNA-based, vector-based, heterologous scheme, any vaccine): 1. completed primary immunization vs. no vaccination (i.e., placebo, no vaccination, or vaccine not directed against COVID-19), 2. booster immunization vs. no vaccination, 3. booster immunization vs. primary immunization. Outcomes of interest were VE against polymerase chain reaction- (PCR) or antigen-test confirmed SARS-CoV-2 infection of “any type” (i.e. studies did not indicate underlying symptoms), “symptomatic COVID-19”, and “severe COVID-19” (including hospitalization, ICU-admission, or death due to SARS-CoV-2 infection). To investigate VE at different time points since vaccination, data was stratified into four time periods (≈14 days, > 14 days up to 3 months, > 3 months up to 6 months, > 6 months). As there was substantial heterogeneity across studies, we abstained from conducting a meta-analysis, but summarized the studies as follows: We assessed the range of VE of any, mRNA-or vector-based vaccines or for heterologous schedules against the outcomes described for the different time strata by including VE estimates from all studies that provide VE estimates for the respective time point (**supplement material, part 4**). If studies reported VE data for more granular time points within the defined time stratum, it was always the estimate of the latest time point within the stratum that contributed to the indicated range (e.g. if studies reported VE after 2-4 and 5-9 weeks after vaccination, the estimate of 5-9 weeks was included in the depicted effect range). Only for the time stratum “≈14 days”, VE estimates closest to 14 days were included. The latter stratum included furthermore VE data that was only assessed at “≥14 days” after primary vaccination, “≥7 days” post booster vaccination or when the time point of assessment after primary immunization was not reported at all. To visualize VE over time after primary and booster vaccination the estimates contributing to the VE range for the respective time category were included in forest plots. Additionally, we assessed the percentage difference of VE over time in studies reporting VE estimates for at least two different time points. We provide the range of observed minimal and maximal differences across studies. For detailed information we refer to the table of all extracted information provided in Appendix 2. ROBINS-I was used to assess risk of bias (3). The certainty of the evidence included in the LSR was rated using the Grading of Recommendations Assessment, Development and Evaluation (GRADE) approach (4, 5).

## 3 Results

### Study screening

We identified a total of 8,428 entries in the database until 14 January 2022. Another 38 potentially relevant studies were added by hand-search until 11 February 2022. After title/abstract and full-text screening, data from 26 studies (6-31) were extracted (**supplement material 1, part 4; figure 1)**. If studies referred to previously published references for further information, these references were considered and information extracted if necessary. For pre-print studies with several versions available, the most recent update published until 11 February 2022 was included.

**Figure 1.**
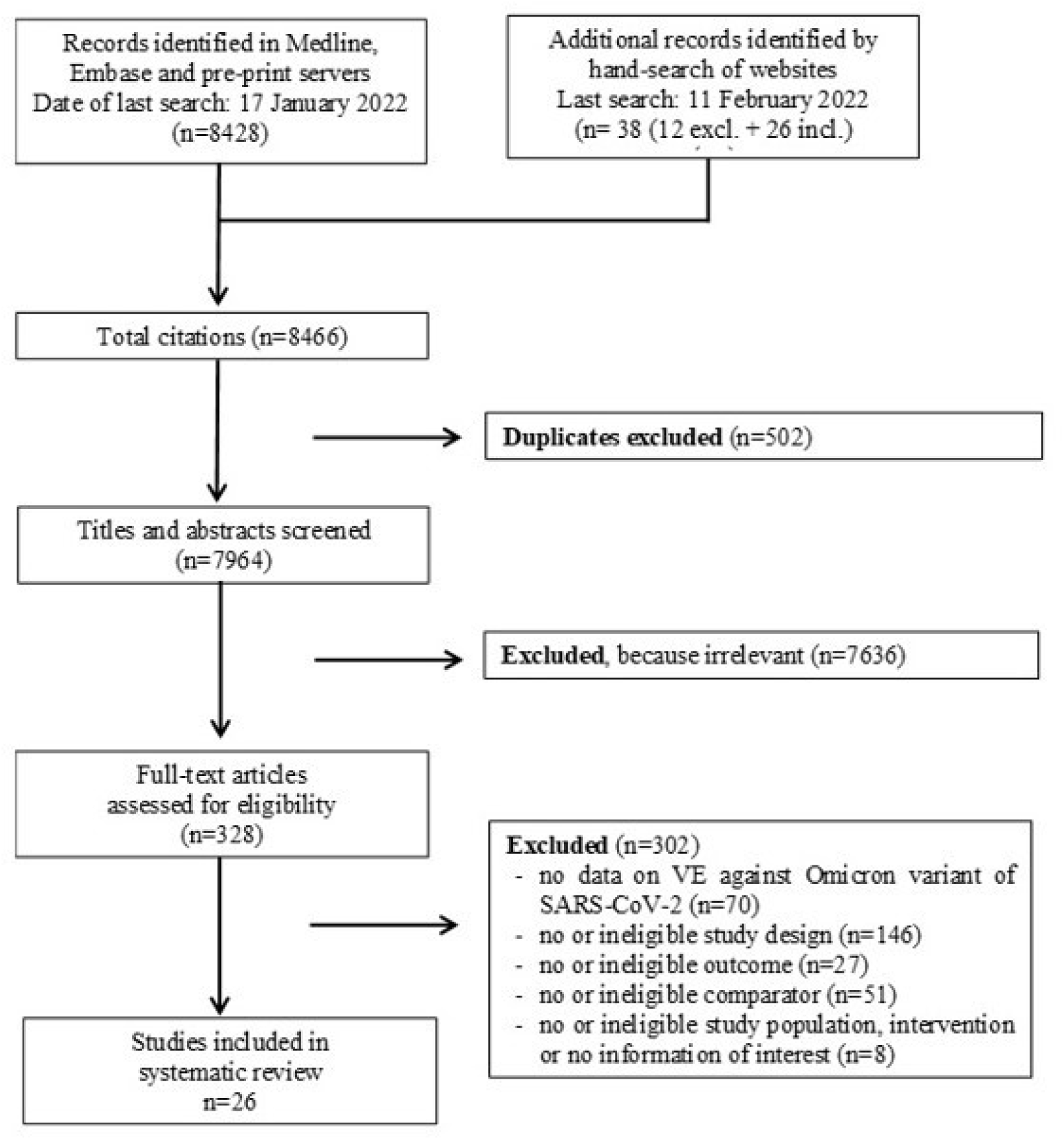
PRISMA flow chart

### Characteristics of included studies

The included studies, of which most (21/26) were not yet peer-reviewed, reported VE estimates against infections (and related outcomes) with only the Omicron variant (5/26) or included VE data against infections with the Delta variant for comparison (21/26). Fourteen studies assessed VE using a test-negative design, eight based on a cohort design, three on a case-control design and one study reported infection rates per vaccination status within a transmission study.

In the studies the SARS-CoV-2 variant causing the investigated outcome was identified either from time periods with known dominant circulation of the Omicron (resp. Delta) variant in the corresponding study location, from whole-genome sequencing (WGS), or through S-gene target failure (SGTF) in PCR assays. Studies using the latter method subdivided the SARS-CoV-2 positive samples by the detection or non-detection of the S-gene in a three gene PCR assay. As the SGTF is characteristic for the Omicron variant, but rare in the Delta variant, studies used the detection of the S-gene as a proxy for the Delta variant, and the “non-detection” as a proxy for the Omicron variant. As most studies report overall VE estimates without further differentiating between WGS- or SGTF-variant assessment the data was synthesized only into “dominance” and “sequencing/SGTF”.

Studies were conducted in ten different countries and mainly used national electronic registries or claims data from the general population for laboratory, immunization and patient characteristics. Only three studies investigated VE for specific populations such as in health care workers, veterans or patients under hemodialysis therapy. Minimum age of included study participants was 12 years (if reported). However, none of the studies provided subgroup data for children or adolescents.

Twenty-two studies reported VE estimates for full primary immunization (as defined per study), 23 for a booster dose (additional dose after full primary immunization). One of the latter studies compared the effect of a second booster dose (fourth doses) with a single booster dose. VE-estimates for several time points after full primary immunization have been reported by eleven studies, eight studies reported VE data for several time points after booster immunization.

Most studies investigated VE of mRNA-based vaccines (13 on Comirnaty, nine on Spikevax), seven studies reported VE for vector-based vaccines (four on Vaxzevria, three on COVID-19 vaccines from Janssen) and seven studies did not differentiate VE per vaccine. None of the studies provided data for Nuvaxovid.

### Prevention of infection with SARS-CoV-2 Omicron variant (without differentiation between asymptomatic or symptomatic cases)

Twelve studies (including between 1,220 and 2.2 million participants) reported the effectiveness of COVID-19 vaccines in preventing infection of any type with SARS-CoV-2 Omicron variant (without differentiation between asymptomatic or symptomatic cases). After full primary immunization VE across all studies ranged between 0 and 62% at “≈14 days” post vaccination, compared to no vaccination. For the time-periods “>14 days up to 3 months”, “>3 months up to 6 months” and “>6 months” after vaccination VE-ranges, assessed across all reported VE estimates, were 4.2 to 42.8%, 0 to 23% and 0 to 8.6%, respectively (**table 1, figure 2**). The data from the two studies reporting VE for at least two time points show a decline of VE between >14 days to up to 6 months by 16 to 34% after vaccination with mRNA-based vaccines (**figure 2**) (7, 14). As there was no VE data over time identified for vector-based vaccines, unspecified vaccine or heterologous schedules respective VE decline could not be assessed.

**Table 1:**
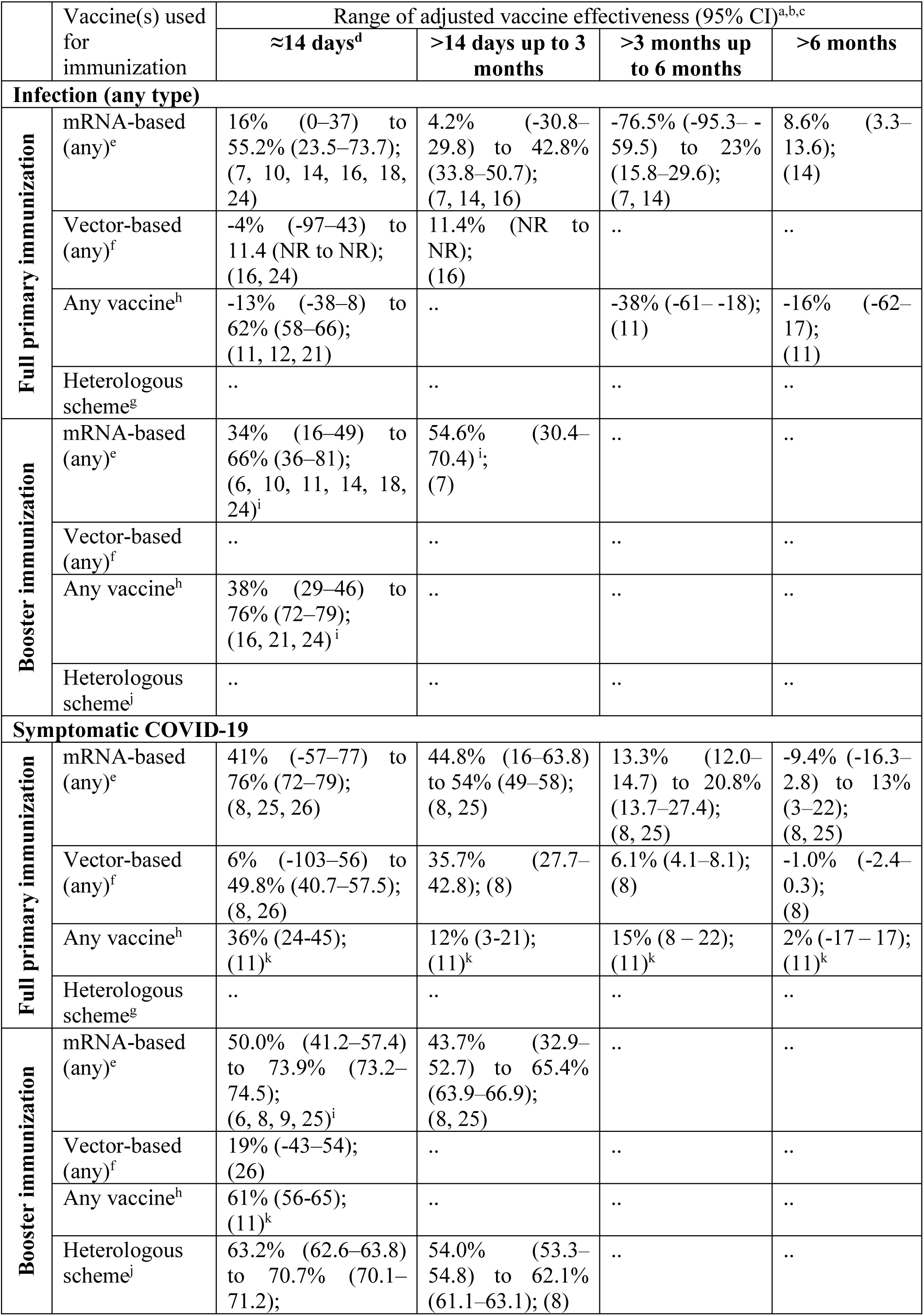
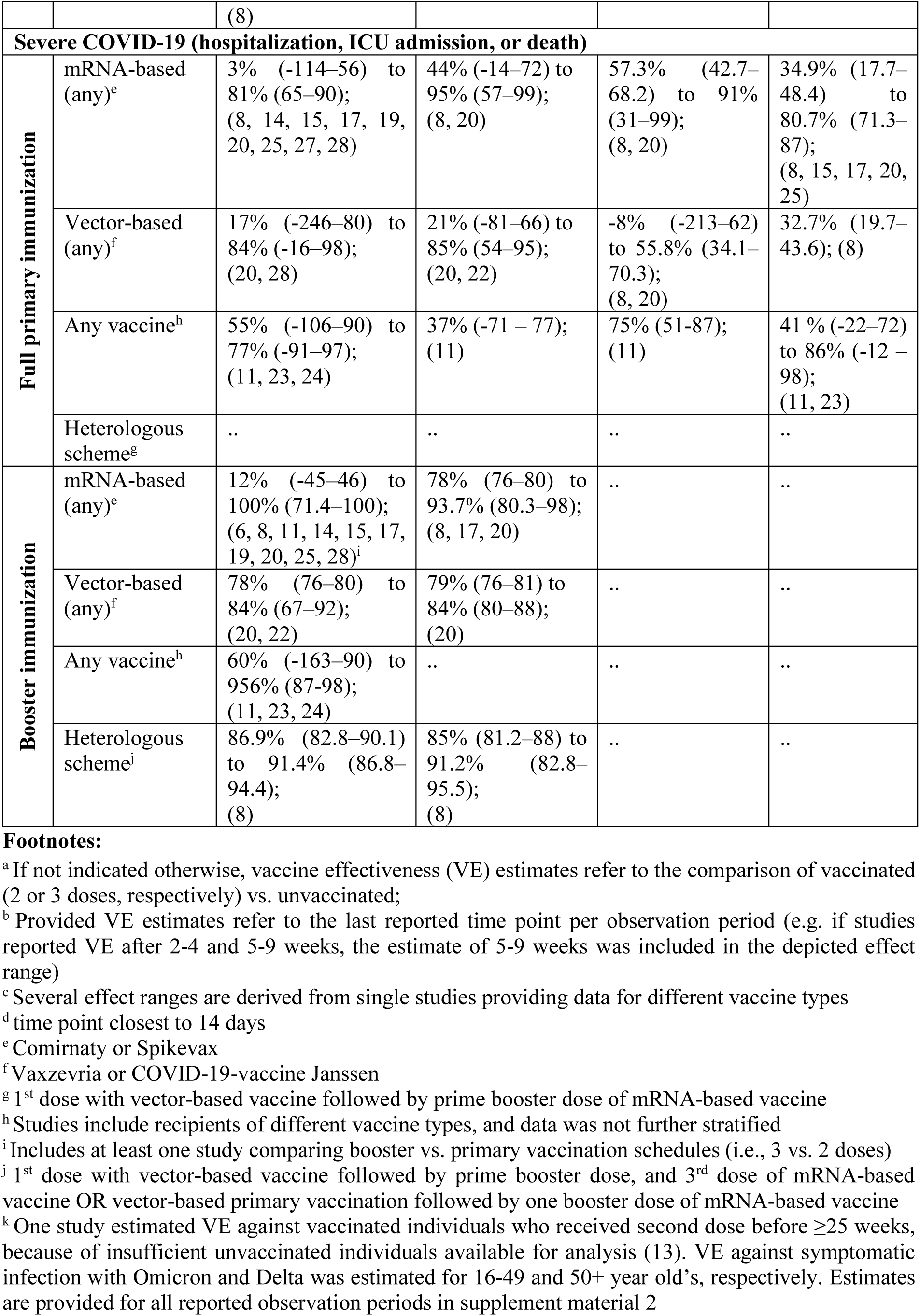
Effectiveness of COVID-19 vaccines against SARS-CoV-2 Omicron variant infection (infection (any type), symptomatic COVID-19, and severe COVID-19 (hospitalization, ICU admission, or death))

| Vaccine(s) used for immunization |  | Range of adjusted vaccine effectiveness (95% CI)<sup>a,b,c</sup> |  |  |  |
| --- | --- | --- | --- | --- | --- |
|  |  | ≈14 days<sup>d</sup> | >14 days up to 3 months | >3 months up to 6 months | >6 months |
| Infection (any type) |  |  |  |  |  |
| Full primary immunization | mRNA-based (any)<sup>e</sup> | 16% (0–37) to 55.2% (23.5–73.7); (7, 10, 14, 16, 18, 24) | 4.2% (-30.8–29.8) to 42.8% (33.8–50.7); (7, 14, 16) | -76.5% (-95.3– -59.5) to 23% (15.8–29.6); (7, 14) | 8.6% (3.3–13.6); (14) |
|  | Vector-based (any)<sup>f</sup> | -4% (-97–43) to 11.4 (NR to NR); (16, 24) | 11.4% (NR to NR); (16) | .. | .. |
|  | Any vaccine<sup>h</sup> | -13% (-38–8) to 62% (58–66); (11, 12, 21) | .. | -38% (-61– -18); (11) | -16% (-62–17); (11) |
|  | Heterologous scheme<sup>g</sup> | .. | .. | .. | .. |
| Booster immunization | mRNA-based (any)<sup>e</sup> | 34% (16–49) to 66% (36–81); (6, 10, 11, 14, 18, 24)<sup>i</sup> | 54.6% (30.4–70.4)<sup>i</sup>; (7) | .. | .. |
|  | Vector-based (any)<sup>f</sup> | .. | .. | .. | .. |
|  | Any vaccine<sup>h</sup> | 38% (29–46) to 76% (72–79); (16, 21, 24)<sup>i</sup> | .. | .. | .. |
|  | Heterologous scheme<sup>j</sup> | .. | .. | .. | .. |
| Symptomatic COVID-19 |  |  |  |  |  |
| Full primary immunization | mRNA-based (any)<sup>e</sup> | 41% (-57–77) to 76% (72–79); (8, 25, 26) | 44.8% (16–63.8) to 54% (49–58); (8, 25) | 13.3% (12.0–14.7) to 20.8% (13.7–27.4); (8, 25) | -9.4% (-16.3–2.8) to 13% (3–22); (8, 25) |
|  | Vector-based (any)<sup>f</sup> | 6% (-103–56) to 49.8% (40.7–57.5); (8, 26) | 35.7% (27.7–42.8); (8) | 6.1% (4.1–8.1); (8) | -1.0% (-2.4–0.3); (8) |
|  | Any vaccine<sup>h</sup> | 36% (24–45); (11)<sup>k</sup> | 12% (3–21); (11)<sup>k</sup> | 15% (8–22); (11)<sup>k</sup> | 2% (-17–17); (11)<sup>k</sup> |
|  | Heterologous scheme<sup>g</sup> | .. | .. | .. | .. |
| Booster immunization | mRNA-based (any)<sup>e</sup> | 50.0% (41.2–57.4) to 73.9% (73.2–74.5); (6, 8, 9, 25)<sup>i</sup> | 43.7% (32.9–52.7) to 65.4% (63.9–66.9); (8, 25) | .. | .. |
|  | Vector-based (any)<sup>f</sup> | 19% (-43–54); (26) | .. | .. | .. |
|  | Any vaccine<sup>h</sup> | 61% (56–65); (11)<sup>k</sup> | .. | .. | .. |
|  | Heterologous scheme<sup>j</sup> | 63.2% (62.6–63.8) to 70.7% (70.1–71.2); | 54.0% (53.3–54.8) to 62.1% (61.1–63.1); (8) | .. | .. |

|  |  | (8) |  |  |  |
| --- | --- | --- | --- | --- | --- |
| **Severe COVID-19 (hospitalization, ICU admission, or death)** |  |  |  |  |  |
| **Full primary immunization** | mRNA-based (any)<sup>e</sup> | 3% (-114–56) to 81% (65–90); (8, 14, 15, 17, 19, 20, 25, 27, 28) | 44% (-14–72) to 95% (57–99); (8, 20) | 57.3% (42.7–68.2) to 91% (31–99); (8, 20) | 34.9% (17.7–48.4) to 80.7% (71.3–87); (8, 15, 17, 20, 25) |
|  | Vector-based (any)<sup>f</sup> | 17% (-246–80) to 84% (-16–98); (20, 28) | 21% (-81–66) to 85% (54–95); (20, 22) | -8% (-213–62) to 55.8% (34.1–70.3); (8, 20) | 32.7% (19.7–43.6); (8) |
|  | Any vaccine<sup>h</sup> | 55% (-106–90) to 77% (-91–97); (11, 23, 24) | 37% (-71 – 77); (11) | 75% (51–87); (11) | 41 % (-22–72) to 86% (-12 – 98); (11, 23) |
|  | Heterologous scheme<sup>g</sup> | .. | .. | .. | .. |
| **Booster immunization** | mRNA-based (any)<sup>e</sup> | 12% (-45–46) to 100% (71.4–100); (6, 8, 11, 14, 15, 17, 19, 20, 25, 28)<sup>i</sup> | 78% (76–80) to 93.7% (80.3–98); (8, 17, 20) | .. | .. |
|  | Vector-based (any)<sup>f</sup> | 78% (76–80) to 84% (67–92); (20, 22) | 79% (76–81) to 84% (80–88); (20) | .. | .. |
|  | Any vaccine<sup>h</sup> | 60% (-163–90) to 956% (87–98); (11, 23, 24) | .. | .. | .. |
|  | Heterologous scheme<sup>j</sup> | 86.9% (82.8–90.1) to 91.4% (86.8–94.4); (8) | 85% (81.2–88) to 91.2% (82.8–95.5); (8) | .. | .. |

<sup>a</sup> If not indicated otherwise, vaccine effectiveness (VE) estimates refer to the comparison of vaccinated (2 or 3 doses, respectively) vs. unvaccinated;
<sup>b</sup> Provided VE estimates refer to the last reported time point per observation period (e.g. if studies reported VE after 2–4 and 5–9 weeks, the estimate of 5–9 weeks was included in the depicted effect range)
<sup>c</sup> Several effect ranges are derived from single studies providing data for different vaccine types
<sup>d</sup> time point closest to 14 days
<sup>e</sup> Comirnaty or Spikevax
<sup>f</sup> Vaxzevria or COVID-19-vaccine Janssen
<sup>g</sup> 1<sup>st</sup> dose with vector-based vaccine followed by prime booster dose of mRNA-based vaccine
<sup>h</sup> Studies include recipients of different vaccine types, and data was not further stratified
<sup>i</sup> Includes at least one study comparing booster vs. primary vaccination schedules (i.e., 3 vs. 2 doses)

**Figure 2.**
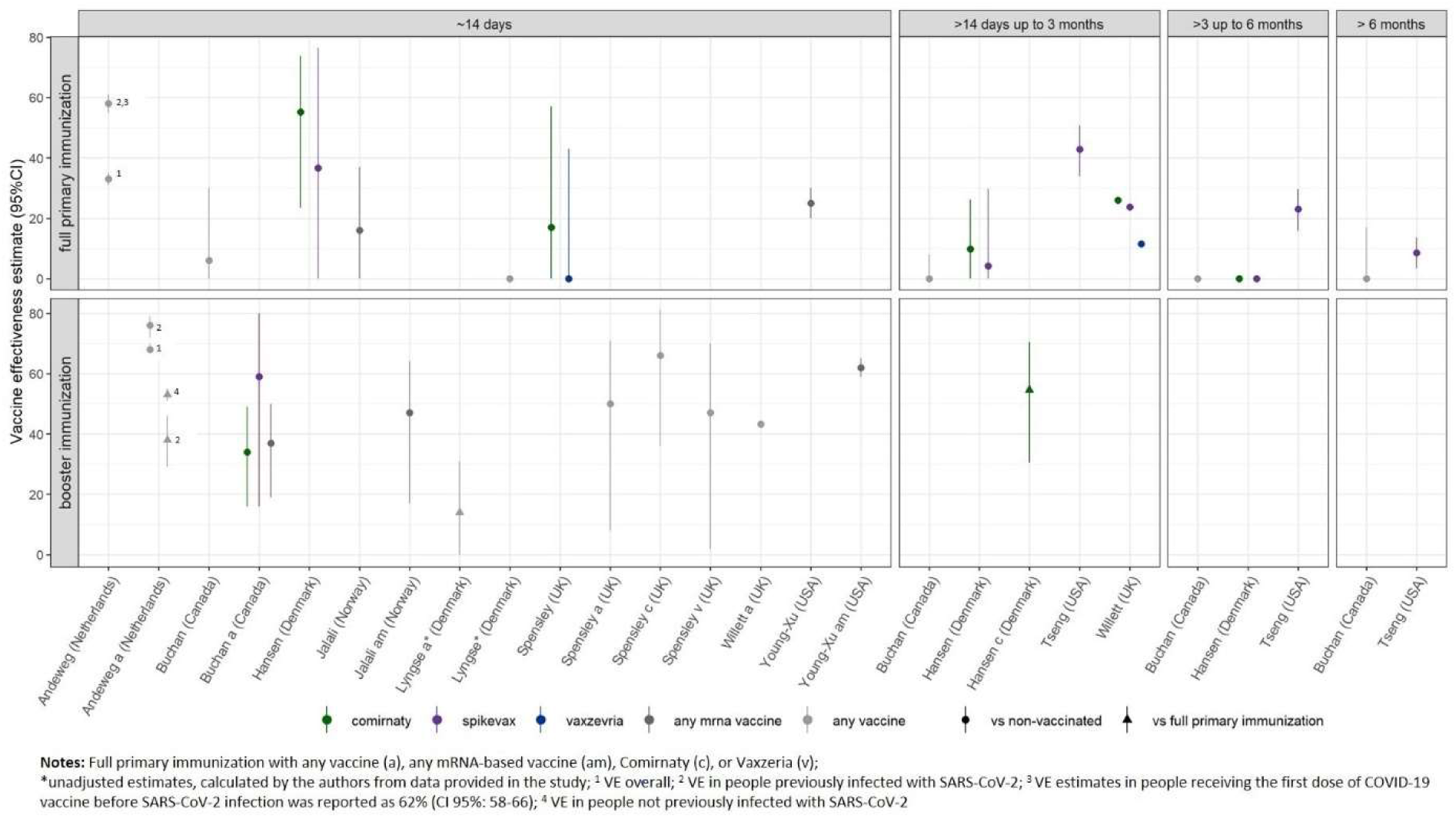
Forest plots of VE estimates against any type of SARS-CoV-2 infection of the Omicron variant after full primary immunization and booster dose, as reported in the study for the defined time strata after immunization. For booster immunization the COVID-19 vaccine used for primary immunization is indicated.

After booster vaccination, VE estimates at “≈14 days” post vaccination across all studies ranged between 34 and 76%, compared to no vaccination, and between 14 and 53%, compared to full primary immunization (12, 21). Follow-up data were insufficient to evaluate waning of immunity after booster vaccination.

The study investigating VE of four vs. three doses of mRNA-based vaccines reported VE estimate of 47% (95% CI 44-47) against Omicron infection at 12 or more days after the fourth dose (29). VE ranges against infection with the Delta variant for the different time points are provided in **supplement material 1, part 5**.

Risk of bias was serious to critical for all assessed studies (see **figure 3**). Key concern was no or insufficient adjustment for confounders.

**Figure 3.**
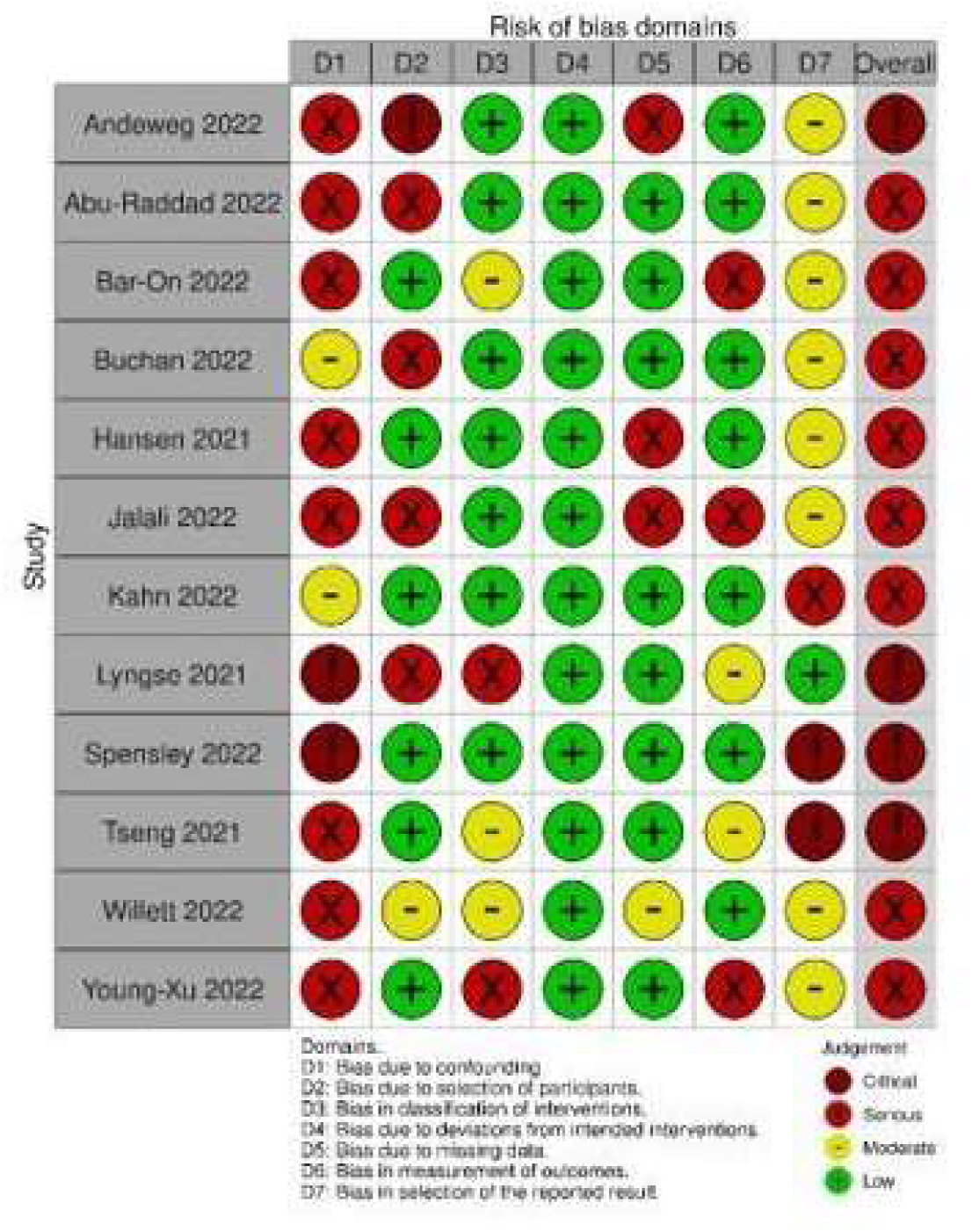
Risk of Bias assessment for SARS-CoV-2 infection (any type) of the Omicron variant

### Prevention of symptomatic COVID-19

Seven studies, including between 430 and 2.2 million participants, estimated the effectiveness of COVID-19 vaccines in preventing symptomatic infection with the SARS-CoV-2 Omicron variant (**supplement material 1, part 4**). Compared to unvaccinated individuals, VE at ≈14 days after vaccination summarized from all studies reporting on this time point ranged between 6 and 76%. For the time-period “>14 days up to 3 months”, “>3 months up to 6 months” and “>6 months” VE ranges were 12-54%, 6.1-20.8%, and 0-13%, respectively. VE estimates from studies that report data for at least two time points suggest a decrease in protective immunity against symptomatic COVID-19 between 45-63% for mRNA-based vaccine recipients, and of 50% for vector-based vaccine recipients over the time period >14 days up to >6 months after vaccination (**figure 4**, (8, 25)).

**Figure 4.**
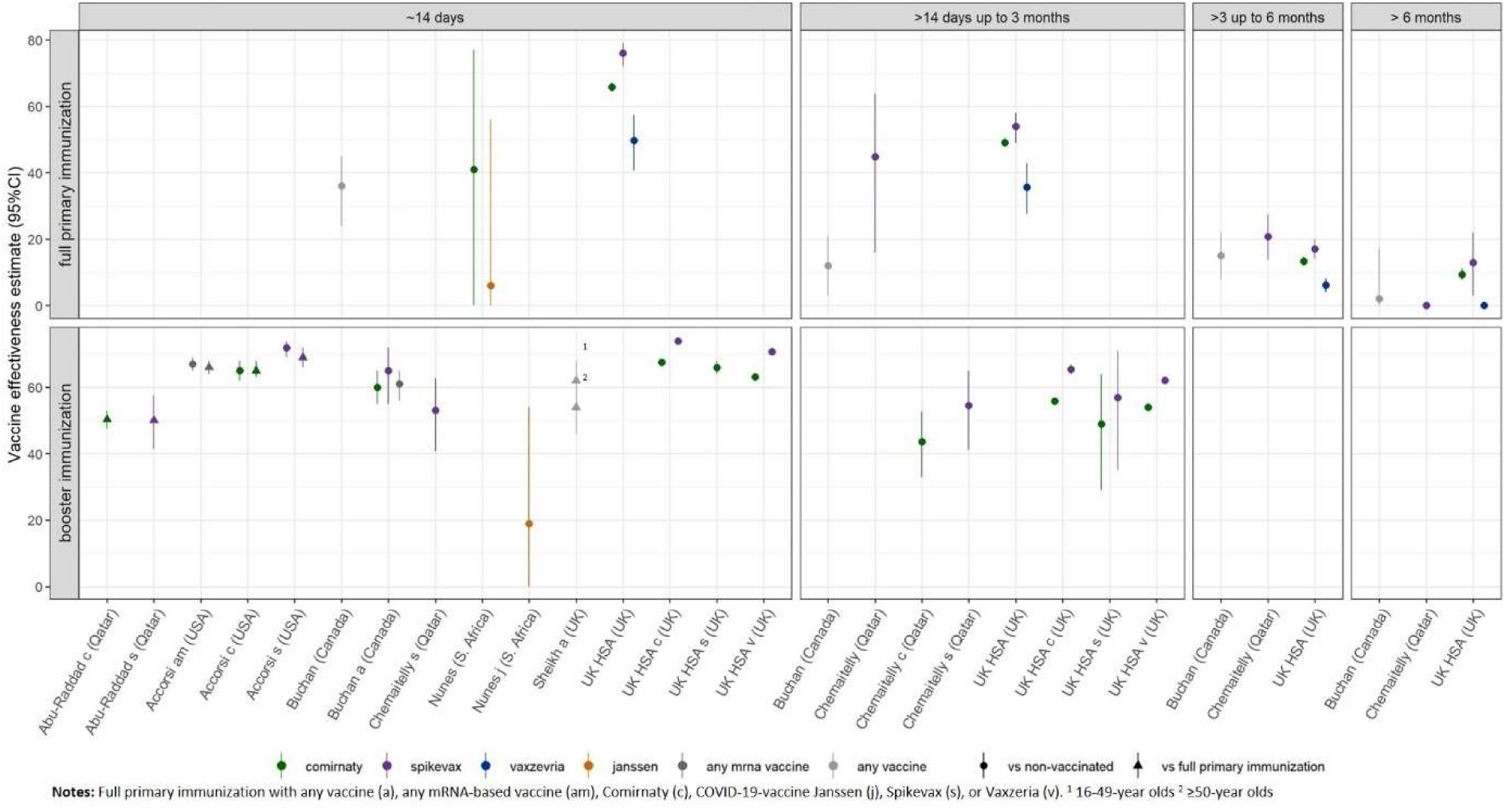
Forest plots of VE estimates against symptomatic COVID-19 due to SARS-CoV-2 infection of the Omicron variant after full primary immunization and booster dose, as reported in the study for the defined time strata after immunization. For booster immunization the COVID-19 vaccine used for primary immunization is indicated.

After booster vaccination, VE against symptomatic infection ranged between 19 and 73.9% at “≈14 days” post vaccination, compared to no vaccination, and between 50 to 68%, compared to primary vaccination (6, 9). For the time point “>14 days up to 3 months” estimates were only reported for the comparison with no vaccination and ranged between 43.7 and 65.4%. No study reported VE for later observation periods. The two studies that provide data for both these time periods show a 9-28% decrease in VE after booster vaccination with a mRNA-based vaccine and between 32-34% after heterologous vaccination schemes (**figure 4** (8, 25)). However, 95% confidence intervals between observation periods were partially overlapping.

Risk of bias was serious to critical for all but one of the assessed studies (see **figure 5**). The remaining study was rated to have a moderate risk. All factors considered relevant for confounding were taken into account, however residual confounding could not be ruled out.

**Figure 5.**
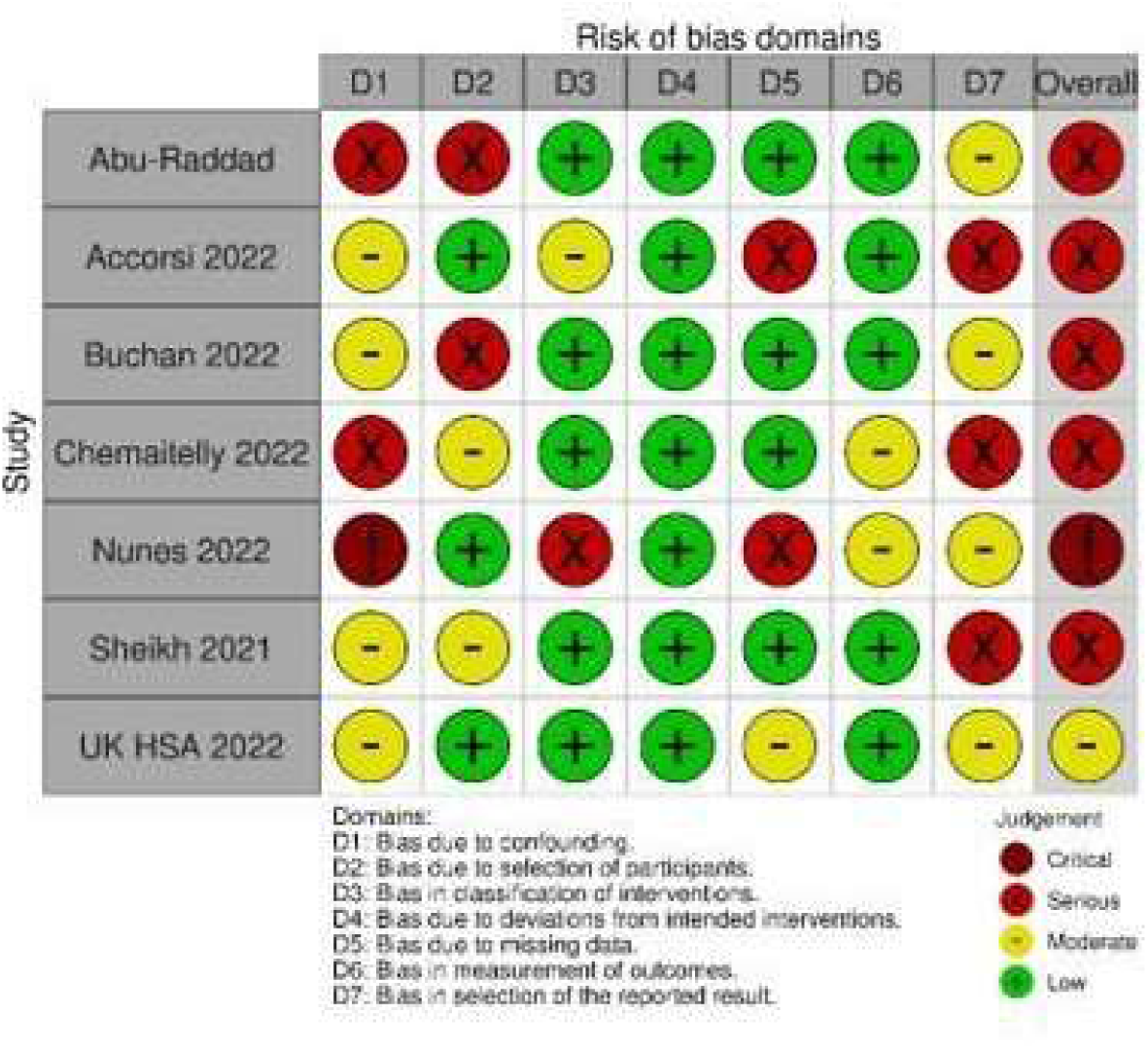
Risk of Bias assessment for symptomatic COVID-19 due to SARS-CoV-2 infection of the Omicron variant

### Prevention of severe COVID-19 (hospitalization, ICU-admission, or death)

VE against severe COVID-19 was assessed in seventeen studies including 1,220 to 2.2 million participants (**supplement material 1, part 4**). VE estimates in primary vaccinated compared to unvaccinated individuals ranged from 3 to 84% at “≈14 days” post vaccination, between 21 and 95% at “>14 days up to 3 months”, between 0 and 91% at “>3 months up to 6 months” and between 32.7 and 86% at “>6 months” after vaccination. Studies reporting VE for at least two time points indicate a decline by up to 40% for mRNA-based vaccines, and 15-67% for vector-based vaccines between 14 days and ≥6 months after vaccination (see **figure 6)** (8, 20). However, 95% confidence intervals were wide and overlapping across time points. One study reported no difference in the first and last time point estimate (17), and another study reported a small non-significant increase (VE at 30-180 days: 73.7% (95% CI 46.8-87); VE at ≥210 days: 80.7% (95% CI 71.3-87); (25)) (**supplement material 2**).

**Figure 6.**
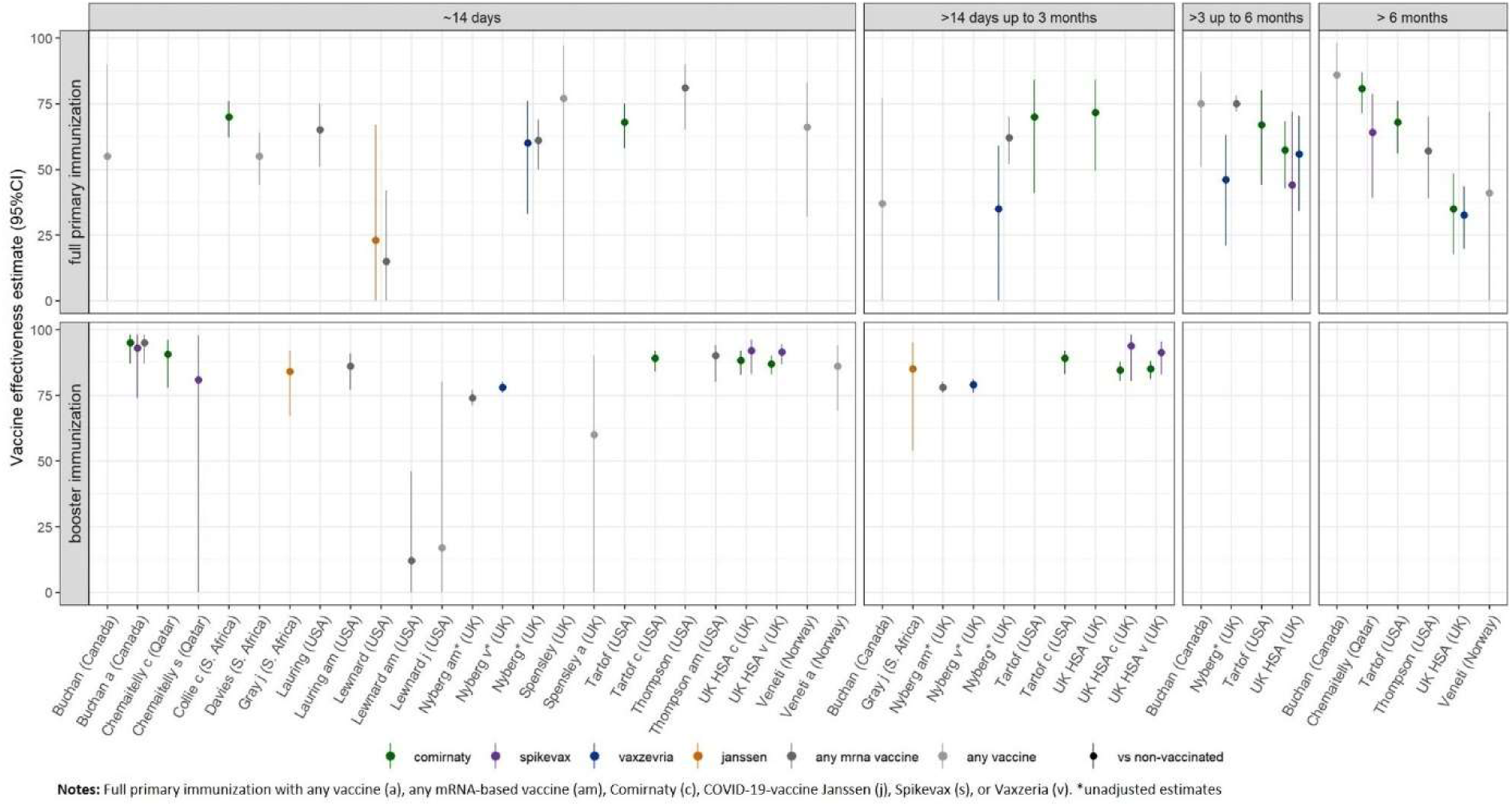
Forest plots of VE estimates against severe COVID-19 (incl. hospitalization, ICU admission or death) due to SARS-CoV-2 infection of the Omicron variant after full primary immunization and booster dose, as reported in the study for the defined time strata after immunization. For booster immunization the COVID-19 vaccine used for primary immunization is indicated.

For booster vaccinated compared to unvaccinated individuals VE ranged from 12 to 100% at “≈14 days” post vaccination and between 78 and 93.7% at “>14 days up to 3 months”. According to three studies, VE after booster vaccination remained stable over this time period (17, 20, 25), irrespective of the vaccine or scheme used (mRNA-, vector-based or heterologous vaccination). Only one study showed a decline of VE of approximately 12% over the respective time period after mRNA-based vaccination (8). VE data was not available for later time periods.

For the comparison against full primary immunization we calculated VE of booster vaccination against severe disease based on data from one study (6) that only reported data for 7 days after booster vaccination (VE: 100% (95% CI 71.4-100)).

The study assessing the effect of a fourth vs. a third mRNA-based vaccine dose reported VE against severe disease of 75% (95% CI 57-86) at ≥12 days after additional booster vaccination (29). Data to assess duration of protection was not available.

In all but one studies risk of bias was serious to critical (see **figure 7**). The remaining study was rated to have a moderate risk due to the potential for residual confounding.

**Figure 7.**
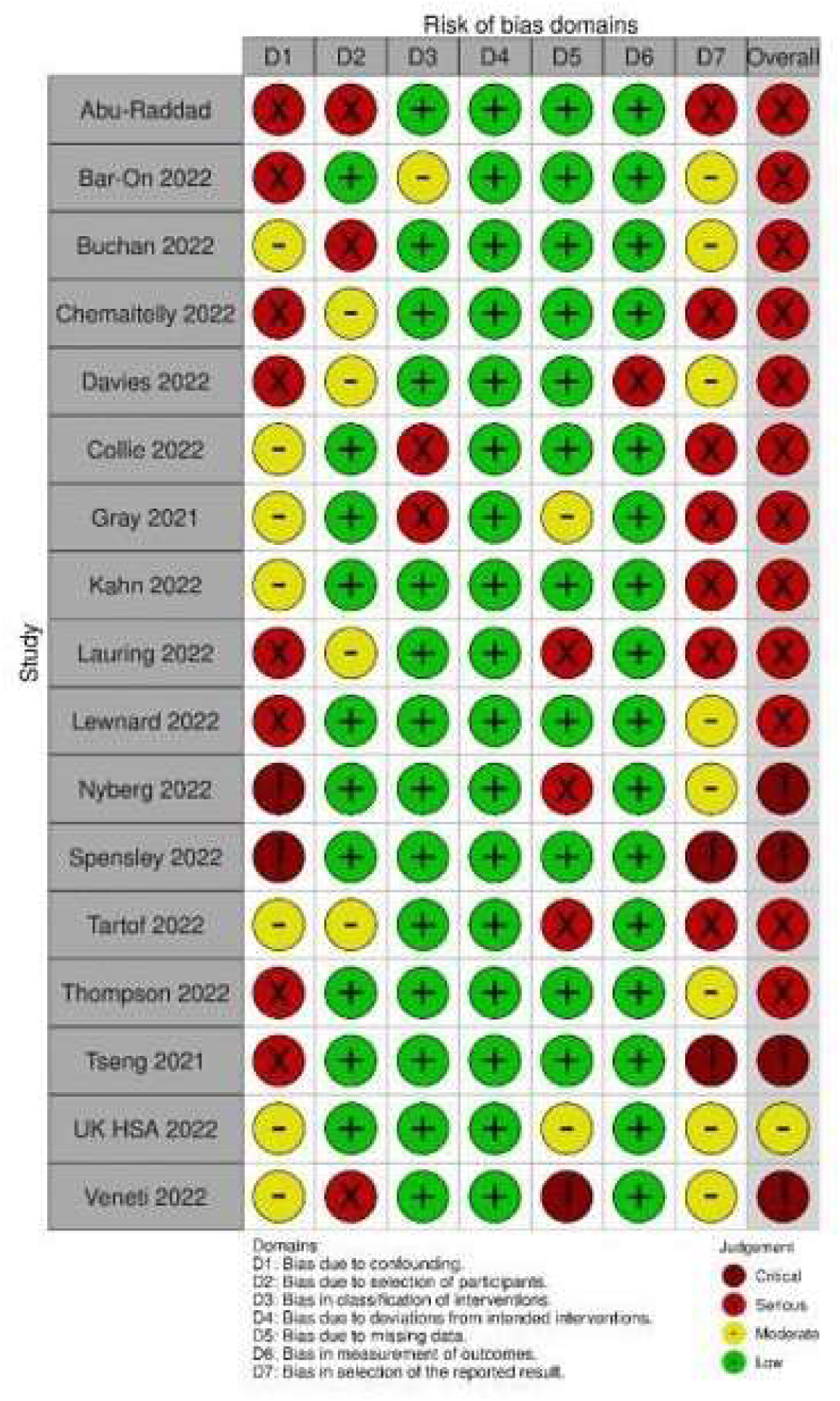
Risk of Bias assessment for severe COVID-19 (incl. hospitalization, ICU admission or death) due to SARS-CoV-2 infection of the Omicron variant

### Effectiveness against the Omicron variant, compared to the Delta variant

Twenty-one studies reported VE estimates both against infections with the Omicron and with the Delta variant (**supplement material 1, part 4; supplement material 1, part 5; supplement material 2**).

In addition to our PICO question, we included eight studies that estimated risk reduction of infections with the Omicron variant compared to those with the Delta variant. Risk ratios from these studies suggest that the risk for any type of SARS-CoV-2 infection and symptomatic COVID-19 despite vaccination is higher for the Omicron variant than for the Delta variant. Instead, risk of severe disease is lower in vaccinated people infected with the Omicron variant compared to those vaccinated and infected with the Delta variant (**supplement material 1, part 6 and 7**).

### Publication bias

Potential publication bias could not be explored through statistical testing and generating funnel plots, as none of the comparisons/outcomes/time points involved sufficient studies.

### GRADE

Overall, GRADE-certainty of evidence is very low for all outcomes, due to the underlying study limitations and serious heterogeneity.

## 4 Discussion

This 3^rd^ update of our LSR provides evidence on VE and duration of protection of EU-approved COVID-19 vaccines against any type of infection, symptomatic infection, and COVID-19 associated severe disease (i.e. hospitalization, ICU admission, or death) caused by the SARS-CoV-2 Omicron variant.

Though evidence is uncertain about the exact level of protection against all investigated outcomes, both after primary and after a booster vaccination, data suggest that VE was higher after booster when compared to primary immunization. Results suggest a rapid decline of vaccine-induced protection after completion of the primary vaccination series. The effect is profound for infections of any type, but less pronounced for severe disease. This is in line with the findings of a recently published meta-regression analysis on pre-Omicron variants (32). VE could be restored to high levels of protection by the booster dose, though first follow-up data do also suggest a waning effect (8, 19, 25). However, VE against severe disease caused by Omicron remained high for at least 3 months post booster immunization (no data for longer follow-up available). Compared to VE against the Delta variant, vaccines were less effective for all reported outcomes.

In the light of the more transmissible Omicron variant and the advancing immunization campaigns across countries, evidence on the need and the timing of booster vaccination is of increasing public health interest. Thus, we decided to adapt our PICO questions and inclusion criteria and considered also studies that compare booster vaccinated individuals with primary vaccinated ones. The effect of waning immunity was assessed through stratifying VE data into multiple observation periods.

As included studies were highly heterogenous (e.g. in terms of analyzed time point after vaccination, study population, applied vaccine schedules, if reported at all) and had serious to critical risk of bias, we decided not to perform meta-analyses, but provide ranges of reported VE estimates across studies to increase transparency and prevent misinterpretation. As studies assessed VE inconsistently for different time points or periods, a standardized assessment for time points of VE evaluation, as suggested by WHO (33), would facilitate the comparison of evidence across studies and better allow a synthesis of the evidence.

Most studies included here used the test-negative design. This study design was initially introduced to estimate VE against seasonal influenza, and thought to control for differences in seeking medical care (34). In the COVID-19 era, the test-negative studies might be prone to bias caused by specific testing strategies or behaviors at the study location. However, those are mainly not reported in the studies, making it difficult to interpret reported estimates, when individuals are not tested due to underlying symptoms. Most studies did not indicate the vaccination schedule applied for full primary immunization or the booster dose. For immunocompromised people a three-dose primary vaccination schedule is recommended by the WHO Strategic Advisory Group of Experts on Immunization to improve immune response. As this recommendation is implemented in many countries^1^, it is possible that VE estimates for a booster schedule include those that received a 3^rd^ doses within an “optimized” schedule for immunocompromised people.

This 3^rd^ update provides a comprehensive overview of the currently available evidence on VE against infection with the SARS-CoV-2 Omicron variant. We provide an in-depth analysis of the VE estimates extracted from the included studies that were identified following a pre-registered protocol. We conducted thorough risk of bias assessment of the studies, and evaluated the certainty of evidence using the GRADE approach.

Due to the highly dynamic publication landscape in this field, additional studies have been published since our last search that are not captured by this analysis. In fact, we are aware of at least two studies reporting on adolescents, which were published after our data cut (35, 36). As in adults, lower VE against infections and severe disease were observed for the Omicron variant, when compared to the Delta variant. However, data on duration of protection are contradicting. While one study reported a decrease in VE against hospitalizations by 6-19% at more than 5 months after primary immunization (35). The second study did not identify a decrease in VE against any infections but was based only on few events (36). In addition to studies not included as they were published after our final search date, we noticed that some studies updated data on preprint servers after initial publication including data on longer follow-up periods. We therefore cannot exclude that authors revised the available pre-print versions including additional data after we completed data extraction for this update. Further, we cannot exclude a risk of potential bias as real-life observation are based on retrospective analyses which are not systematically registered. Thus, intent and possibility of publication might depend on observed results. However, due to the increase of publications on pre-print servers, the risk of publication bias is probably low.

Additional data limitations stem from the fact that VE estimates for more than one vaccine were reported.

## 5 Conclusion

Current evidence suggests that the effectiveness of COVID-19 vaccines licensed in the EU is low after primary vaccination, and improved after booster vaccination in preventing infections with the SARS-CoV-2 Omicron variant. For both primary and booster immunization it is characterized by a rapid decrease over time. VE against severe courses of COVID-19 remains generally high.

Studies included in this update were very heterogenous, therefore, pooling of estimates was not appropriate. To allow statistical synthesizes of effects, studies need to be comparable for clinical and meta-epidemiological aspects. Thus, certain standards for VE studies, as suggested by WHO, are useful to better inform vaccination guidelines and reliably assess the public health impact of vaccination campaigns.

## Data Availability

All data produced in the present work are contained in the manuscript

## 7 Author Contributions

WKS was first and second reviewer, responsible for data extraction, performed bias assessment and drafted the manuscript. VP was first and second reviewer, responsible for data extraction, performed bias assessment, conducted GRADE assessment and contributed to the manuscript.

AP was first and second reviewer, responsible for data extraction, performed bias assessment and contributed to the manuscript. MB was first reviewer, responsible for data extraction, performed bias assessment and contributed to the manuscript. MW, SK, LSD and BG were first reviewers and contributed to the manuscript. MT provided input into the interpretation of the results and developed forest plots. SVB and JK contributed to the manuscript. OW held general oversight of the conducted work and revised the manuscript. TH conceived the study and contributed to the manuscript. All authors contributed to the interpretation of the data and provided important intellectual content to the manuscript.

## 9 Acknowledgements

We sincerely thank the authors of Andrews et al. and Kahn et al. for providing us with additional data.

## Footnotes

1 An additional dose is recommended for immunocompromised people in all eight countries where included studies that reported VE estimates on booster immunization were conducted (Canada, Denmark, Netherlands, Norway, Qatar, South Africa, United Kingdom, United States of America).

