## Supplement material 1 for "Facing the Omicron variant – How well do vaccines protect against mild and severe COVID-19? Third interim analysis of a living systematic review"

### Part 1: PRISMA checklist

| **Section/topic** | **#** | **Checklist item** | **Reported on page #** |
| --- | --- | --- | --- |
| **TITLE** | | |  |
| Title | 1 | Identify the report as a systematic review, meta-analysis, or both. | 1 |
| **ABSTRACT** | | |  |
| Structured summary | 2 | Provide a structured summary including, as applicable: background; objectives; data sources; study eligibility criteria, participants, and interventions; study appraisal and synthesis methods; results; limitations; conclusions and implications of key findings; systematic review registration number. | 1 and 2 |
| **INTRODUCTION** | | |  |
| Rationale | 3 | Describe the rationale for the review in the context of what is already known. | {, 2016 #507}3 |
| Objectives | 4 | Provide an explicit statement of questions being addressed with reference to participants, interventions, comparisons, outcomes, and study design (PICOS). | 3 and Supplement material 1, part 2 |
| **METHODS** | | |  |
| Protocol and registration | 5 | Indicate if a review protocol exists, if and where it can be accessed (e.g., Web address), and, if available, provide registration information including registration number. | 3 |
| Eligibility criteria | 6 | Specify study characteristics (e.g., PICOS, length of follow-up) and report characteristics (e.g., years considered, language, publication status) used as criteria for eligibility, giving rationale. | 3 |
| Information sources | 7 | Describe all information sources (e.g., databases with dates of coverage, contact with study authors to identify additional studies) in the search and date last searched. | 3 and Supplement material 1, part 3 |
| Search | 8 | Present full electronic search strategy for at least one database, including any limits used, such that it could be repeated. | Supplement material 1, part 3 |
| Study selection | 9 | State the process for selecting studies (i.e., screening, eligibility, included in systematic review, and, if applicable, included in the meta-analysis). | 3 and 4 |
| **Section/topic** | **#** | **Checklist item** | **Reported on page #** |
| Data collection process | 10 | Describe method of data extraction from reports (e.g., piloted forms, independently, in duplicate) and any processes for obtaining and confirming data from investigators. | 3 |
| Data items | 11 | List and define all variables for which data were sought (e.g., PICOS, funding sources) and any assumptions and simplifications made. | 3 and 4 |
| Risk of bias in individual studies | 12 | Describe methods used for assessing risk of bias of individual studies (including specification of whether this was done at the study or outcome level), and how this information is to be used in any data synthesis. | 4 |
| Summary measures | 13 | State the principal summary measures (e.g., risk ratio, difference in means). | 3 and 4 |
| Synthesis of results | 14 | Describe the methods of handling data and combining results of studies, if done, including measures of consistency (e.g., I^2^) for each meta-analysis. | 3 and 4 |
| Risk of bias across studies | 15 | Specify any assessment of risk of bias that may affect the cumulative evidence (e.g., publication bias, selective reporting within studies). | 7 |
| Additional analyses | 16 | Describe methods of additional analyses (e.g., sensitivity or subgroup analyses, meta-regression), if done, indicating which were pre-specified. | Not applicable |
| **RESULTS** | | | |
| Study selection | 17 | Give numbers of studies screened, assessed for eligibility, and included in the review, with reasons for exclusions at each stage, ideally with a flow diagram. | 4, 5 and Figure 1 |
| Study characteristics | 18 | For each study, present characteristics for which data were extracted (e.g., study size, PICOS, follow-up period) and provide the citations. | Supplement material, part 4 |
| Risk of bias within studies | 19 | Present data on risk of bias of each study and, if available, any outcome level assessment (see item 12). | Figure 3, Figure 5, Figure 7 |
| Results of individual studies | 20 | For all outcomes considered (benefits or harms), present, for each study: (a) simple summary data for each intervention group (b) effect estimates and confidence intervals, ideally with a forest plot. | Figure 2; Figure 5, Figure 7, Supplement material 2 |
| **Section/topic** | **#** | **Checklist item** | **Reported on page #** |
| Synthesis of results | 21 | Present results of each meta-analysis done, including confidence intervals and measures of consistency. | 5-7, Table 1, Supple-ment material 1, part 5 and 7 |
| Risk of bias across studies | 22 | Present results of any assessment of risk of bias across studies (see Item 15). | Not applicable |
| Additional analysis | 23 | Give results of additional analyses, if done (e.g., sensitivity or subgroup analyses, meta-regression [see Item 16]). | Not applicable |
| **DISCUSSION** | | |  |
| Summary of evidence | 24 | Summarize the main findings including the strength of evidence for each main outcome; consider their relevance to key groups (e.g., healthcare providers, users, and policy makers). | 7-9 |
| Limitations | 25 | Discuss limitations at study and outcome level (e.g., risk of bias), and at review-level (e.g., incomplete retrieval of identified research, reporting bias). | 8-9 |
| Conclusions | 26 | Provide a general interpretation of the results in the context of other evidence, and implications for future research. | 9 |
| **FUNDING** | | |  |
| Funding | 27 | Describe sources of funding for the systematic review and other support (e.g., supply of data); role of funders for the systematic review. | 9 |

*From:* Moher D, Liberati A, Tetzlaff J, Altman DG, The PRISMA Group (2009). Preferred Reporting Items for Systematic Reviews and Meta-Analyses: The PRISMA Statement. PLoS Med 6(7): e1000097. doi:10.1371/journal.pmed1000097

### Part 2: PICO question

**Participants/population:** Male and female participants of age groups ≥ 12 years of age.

**Intervention:** Any vaccine against COVID-19 which has been approved for use in the European Union (or will be approved soon), including complete dosing schedules, i.e. full primary vaccination series and/or booster series.

**Comparators/control:** placebo, no vaccination or a vaccine not directed against COVID-19 (active comparator), but also including head-to-head trials directly comparing different vaccines against COVID-19 and those comparing full vaccination series with booster series.

**Outcomes:** 1. Efficacy and effectiveness-related outcomes: SARS-CoV-2 infection (PCR-confirmed); hospitalization due to COVID-19 (PCR-confirmed); ICU admission due to COVID-19 (PCR-confirmed); intubation and oxygen supply due to COVID-19 (PCR-confirmed); death due to COVID-19 (PCR-confirmed).
2. Safety-related outcomes: local reactions; systemic events; severe adverse events; enhanced COVID-19 disease; adverse events of special interest (AESI), including solicited and unsolicited events

**Setting (for the current analysis):** (Dominant circulation of the) Omicron variant of SARS-CoV-2.

### Part 3: Search strategy

The following searches will be combined with the terms "vaccin*" and "immuniz*" and the brand names of the approved vaccines.*

#### Search Syntax PubMed 1:

("Severe Acute Respiratory Syndrome Coronavirus 2" [Supplementary Concept] OR "COVID-19" [Supplementary Concept] OR "COVID 19 diagnostic testing" [Supplementary Concept] OR "COVID 19 drug treatment" [Supplementary Concept] OR "COVID 19 serotherapy"[Supplementary Concept] OR "COVID 19 vaccine" [Supplementary Concept] OR "Severe Acute Respiratory Syndrome Coronavirus 2"[tiab] OR ncov*[tiab] OR COVID*[tiab] OR sars-cov-2[tiab] OR "sars cov 2"[tiab] OR "SARS Coronavirus 2"[tiab] OR "Severe Acute Respiratory Syndrome CoV 2"[tiab] OR "Wuhan coronavirus"[tiab] OR "Wuhan seafood market pneumonia virus"[tiab] OR "SARS2"[tiab] OR "2019-nCoV"[tiab] OR "hcov-19"[tiab] OR „novel 2019 coronavirus“[tiab] OR "2019 novel coronavirus*"[tiab] OR „novel coronavirus 2019*“[tiab] OR "2019 novel human coronavirus*"[tiab] OR „human coronavirus 2019“[tiab] OR "coronavirus disease-19"[tiab] OR "corona virus disease-19"[tiab] OR "coronavirus disease 2019"[tiab] OR "corona virus disease 2019"[tiab] OR "2019 coronavirus disease"[tiab] OR "2019 corona virus disease"[tiab] OR „novel coronavirus disease 2019“[tiab] OR „novel coronavirus infection 2019“[tiab] OR "new coronavirus*"[tiab] OR "coronavirus outbreak"[tiab] OR "coronavirus epidemic"[tiab] OR "coronavirus pandemic"[tiab] OR "pandemic of coronavirus"[tiab]) AND ("2019/12/01"[PDAT] : "2099/12/31"[PDAT])

#### Search Syntax PubMed 2:

("wuhan"[tiab] or china[tiab] or hubei[tiab]) AND ("Severe Acute Respiratory Syndrome Coronavirus 2"[Supplementary Concept] OR "COVID-19" [Supplementary Concept] OR "COVID 19 diagnostic testing"[Supplementary Concept] OR "COVID 19 drug treatment"[Supplementary Concept] OR "COVID 19 serotherapy"[Supplementary Concept] OR "COVID 19 vaccine"[Supplementary Concept] OR "coronavirus*"[tiab] OR "corona virus*"[tiab] OR ncov[tiab] OR COVID*[tiab] OR sars*[tiab])

#### Search Syntax Embase 1:

('severe acute respiratory syndrome coronavirus 2':ti,ab OR 'severe acute respiratory syndrome coronavirus 2'/exp OR 'COVID 19'/exp OR ncov*:ti,ab OR COVID*:ti,ab OR 'sars cov 2':ti,ab OR 'sars-cov-2':ti,ab OR 'sars coronavirus 2':ti,ab OR 'sars coronavirus 2'/exp OR 'severe acute respiratory syndrome cov 2':ti,ab OR 'wuhan coronavirus':ti,ab OR 'wuhan seafood market pneumonia virus':ti,ab OR sars2:ti,ab OR '2019-ncov':ti,ab OR 'hcov-19':ti,ab OR 'novel 2019 coronavirus':ti,ab OR '2019 novel coronavirus*':ti,ab OR 'novel coronavirus 2019'/exp OR '2019 novel human coronavirus*':ti,ab OR 'human coronavirus 2019':ti,ab OR 'coronavirus disease-19':ti,ab OR 'corona virus disease-19':ti,ab OR 'coronavirus disease 2019':ti,ab OR 'coronavirus disease 2019'/exp OR 'corona virus disease 2019':ti,ab OR '2019 coronavirus disease':ti,ab OR 'novel coronavirus 2019*':ti,ab OR 'novel coronavirus disease 2019':ti,ab OR 'novel coronavirus infection 2019':ti,ab OR '2019 corona virus disease':ti,ab OR 'new coronavirus*':ti,ab OR 'coronavirus outbreak':ti,ab OR 'coronavirus epidemic':ti,ab OR 'coronavirus pandemic':ti,ab OR 'pandemic of coronavirus':ti,ab OR 'severe acute respiratory syndrome coronavirus 2 vaccine'/exp OR 'COVID 19 vaccine'/exp) AND 2020:py

#### Search Syntax Embase 2:

(wuhan:ti,ab OR china:ti,ab OR hubei:ti,ab) AND ('severe acute respiratory syndrome coronavirus 2':ti,ab OR 'severe acute respiratory syndrome coronavirus 2'/exp OR 'severe acute respiratory syndrome coronavirus 2' OR 'COVID*':ti,ab OR 'COVID 19'/exp OR 'COVID 19' OR coronavirus*:ti,ab OR 'corona virus*':ti,ab OR ncov:ti,ab OR COVID*:ti,ab OR sars*:ti,ab OR 'sars coronavirus 2'/exp)

**Manual search in ArRvix, BioRvix, ChemRvix, MedRvix, Preprints.org, ResearchSquare und SSRN:**

Manual search at Websites of European Centre for Disease Prevention and Control (ECDC), US Centers for Disease Control, Public Health Agency of Canada, Public Health England, Hauté Autorite´de Santé (France), testimonicos <https://app.iloveevidence.com/loves/5e6fdb9669c00e4ac072701d?utm=ile>

and the view-hub from the International Vaccine Access Center provided by John Hopkins Bloomberg School of Public Health

### Part 4: Table 1: Characteristics of studies included for data extraction

| Author, year of publication | Study location | Study design | Study period | Study population, n [characteristics] | Variant analysed [method used to identify variant] | Vaccine analysed | Vaccine schedule analysed | Outcome analysed [definition of severe COVID-19 applied] | Funding received | Conflict of interest declared |
| --- | --- | --- | --- | --- | --- | --- | --- | --- | --- | --- |
| Abu-Raddad, 2022 (preprint) (1) | Qatar | cohort study | 05.01.2021 - 09.01.2022 | general population, n=2183465  [median age: 43 (IQR, 35-53), not previously infected] | Omicron (B.1.1.529), Delta (B.1.617.2) [dominance] | Comirnaty, Spikevax | booster immunization [not reported] | infection (any type), symptomatic COVID-19,  severe COVID-19 [hospitalization and death] | not reported | not declared |
| Accorsi, 2022 (2) | United States of America | test-negative | 10.12.2021 - 01.01.2022 | general population, n=70155  [mean age: 40,3 (SD ±15,6)] | Omicron (B.1.1.529), Delta (B.1.617.2) [sequencing/s-gene target failure] | Comirnaty, Spikevax | booster immunization [homologous and heterologous] | symptomatic COVID-19 | Centers for Disease Control and Prevention | not declared |
| Andeweg, 2022 (preprint) (3) | Netherlands | test-negative | 22.11.2021 - 19.01.2022 | general population, n=528488 tests administered [adults, differentiation between previously infected and not previously infected] | Omicron (B.1.1.529), Delta (B.1.617.2) [sequencing/s-gene target failure] | any vaccine | full primary immunization [homologous], booster immunization [not reported] | infection (any type) | not reported | not reported |
| Bar-On, 2022 (preprint) (4) | Israel | cohort study | 15.01.2022 - 27.01.2022 (for infection)  15.01.2022 - 21.01.2022 (for severe disease) | general population, n=1138681  [≥60 years of age] | Omicron (B.1.1.529) [dominance] | Comirnaty | 2. booster immunization [homologous] | infection (any type), severe COVID-19  [“resting respiratory rate >30 breaths/ minute, an oxygen saturation <94% while breathing ambient air, or a ratio of partial pressure of arterial oxygen to fraction of inspired oxygen < 300).”] | no funding received | not declared |
| Buchan, 2022 (preprint) (5) | Canada | test-negative | 22.11.2021 - 19.12.2021 | general population, n=134435  [adults, test-positive: mean 34,87 (SD ±13,71); test-negative: mean 45.04 (SD ±17.66), including previously infected] | Omicron (B.1.1.529) [sequencing/s-gene target failure], Delta (B.1.617.2) [dominance] | any vaccine | full primary immunization, booster immunization [not reported] | infection (any type), symptomatic COVID-19, severe COVID-19 [hospitalization or death] | Canadian Immunization Research Network, Public Health Agency of Canada, ICES | yes |
| Chemaitelly, 2022 (preprint) (6) | Qatar | case-control | 23.12.2021 - 02.02.2022 | general population, n=105504  [median age of cases: 12 (IQR 6-32) (18 (IQR 6-34) for booster dose comirnaty); median age of controls: 12 (IQR 5-32) (19 (IQR 5-34) for booster dose comirnaty)] | Omicron (B.1.1.529) [sequencing/s-gene target failure] | Comirnaty, Spikevax | full primary immunization, booster immunization [homologous] | symptomatic COVID-19, severe COVID-19 [acute-care hospitalizations, ICU hospitalizations, or fatal COVID-19] | Gilead Sciences | yes |
| Collie, 2021 (7) | South Africa | test-negative | 15.11.2021 - 07.12.2021 (Omicron period) 01.09.2021 - 30.10.2021 (Delta period) | general population, n= 211610  [including previously infected] | Omicron (B.1.1.529), Delta (B.1.617.2) [dominance] | Comirnaty | full primary immunization [homologous] | severe COVID-19 [hospitalization for Covid-19 based on admission data] | not reported | not reported |
| Davies, 2022 (preprint) (8) | South Africa | cohort study | 14.11.2021 - 11.12.2021 | general population, n=9499 (5144 during Omicron period; 4355 during Delta period)  [64% 20-39 year olds; 17% 40-49 year olds; 11% 50-59 year olds; 5% 60-69 year olds, 2,7% ≥70 year olds, including previously infected] | Omicron (B.1.1.529), Delta (B.1.617.2) [dominance] | Comirnaty, Janssen | full primary immunization [not reported] | severe COVID-19 [“Hospitalization included admission within 14 days before or after a COVID-19 diagnosis except for admissions to long term psychiatric or rehabilitation facilities”] | Western Cape Provincial Health Data Centre from the Western Cape Department of Health, the US National Institutes for Health, the Bill and Melinda Gates Foundation, the United States Agency for International Development, the European Union, the Wellcome Trust, the Medical Research Council of South Africa | none declared |
| Gray, 2021 (preprint) (9) | South Africa | test-negative | 08.11.2021 - 17.12.2021 | health care workers, n=69092  [including previously infected] | Omicron (B.1.1.529) [dominance] | Janssen | booster immunization [homologous] | severe COVID-19 [COVID19 [hospital] admission for Omicron] | National Treasury of South Africa, the National Department of Health, Solidarity Response Fund NPC, The Michael & Susan Dell Foundation, The Elma Vaccines and Immunization Foundation Grant, the Bill & Melinda Gates Foundation | not declared |
| Hansen, 2021 (preprint) (10) | Denmark | cohort study (time-to-event analysis) | 20.11.2021 - 12.12.2021 | general population, n= not reported  [≥12 year olds, median age: 28 years, not previously infected] | Omicron (B.1.1.529) [sequencing/s-gene target failure], Delta (B.1.617.2) [dominance] | Comirnaty, Spikevax | full primary immunization [homologous] | infection (any type) | not reported | not reported |
| Jalali, 2022 (preprint) (11) | Norway | cohort study | 14.12.2021 - 14.01.2022 | houshold members, n=1470  [≥16 year olds, not previously infected] | Omicron (B.1.1.529), Delta (B.1.617.2) [sequencing/s-gene target failure] | Comirnaty, Spikevax | full primary immunization, booster immunization [not reported] | infection (any type) | Norwegian Institute of Public Health | not declared |
| Kahn, 2022 (preprint) (12) | Sweden | test-negative | 27.12.2021 - 09.01.2022 | general population, n=55269  [including previously infected] | Omicron (B.1.1.529), Delta (B.1.617.2) [dominance] | any vaccine | full primary immunization, booster immunization [not reported] | infection (any type), severe COVID-19  [“at least 24 hour-hospitalization five days before until 14 days after a positive test and with a need of oxygen supply ≥ 5 L/min or admittance to an intensive care unit”] | Swedish Research Council, Sweden’s Innovation Agency,  internal grants for thematic collaboration initiatives at Lund University | not declared |
| Lauring, 2022 (preprint) (13) | United States of America | case-control | 11.03.2021 - 14.01.2022 | hospitalized patients, n=10472  [median age of cases: 57 (IQR 43-69); median age of controls: 63 (IQR 50-72)] | Omicron (B.1.1.529), Delta (B.1.617.2) [sequencing/s-gene target failure] | Comirnaty, Spikevax | full primary immunization, booster immunization [not reported] | severe COVID-19 ["hospitalized with a clinical syndrome consistent with acute COVID-19 […] and a positive molecular or antigen test for SARS-CoV-2 within 10 days of symptom onset."] | US CDC and Science Award from the National Center for Advancing Translational Sciences, National Institutes of Health | yes |
| Lewnard, 2022 (preprint) (14) | United States of America | cohort study | 30.11.2021 - 01.01.2022 | general population, n=69062  [including previously infected] | Omicron (B.1.1.529), Delta (B.1.617.2) [sequencing/s-gene target failure] | Comirnaty, Janssen, Spikevax, | full primary immunization, booster immunization [homologous and heterologous] | severe COVID-19 [symptomatic hospital admission] | US Centers for Disease Control and Prevention | not reported |
| Lyngse, 2021 (preprint) (15) | Denmark | other | 09.12.2021 - 19.12.2021 | houshold members, n=27874  [including previously infected] | Omicron (B.1.1.529), Delta (B.1.617.2) [sequencing/s-gene target failure] | any vaccine | full primary immunization, booster immunization [not reported] | infection (any type) | Independent Research Fund Denmark, Novo Nordisk Foundation, the Danish National Research Foundation, Statens Serum Institut | yes |
| Nunes, 2022 (preprint) (16) | South Africa | test-negative | 24.11.2021 - 21.12.2021 | health care workers, n=433  [differentiating between previously infect] | Omicron (B.1.1.529) [sequencing/s-gene target failure] | Janssen | full primary immunization, booster immunization [homologous] | symptomatic COVID-19 | European & Developing Countries Clinical Trials Partnership; The Bill & Melinda Gates Foundation; Department of Science and Technology and National Research Foundation: South African Research Chair Initiative in Vaccine Preventable Diseases; the South African Medical Research Council | yes |
| Nyberg, 2022 (preprint) (17) | United Kingdom | cohort study | 22.11.2021 - 09.01.2022 | general population, n=1516533 | Omicron (B.1.1.529), Delta (B.1.617.2) [sequencing/s-gene target failure] | Comirnaty, Spiekvax, Vaxzevria | full primary immunization, booster immunization [homologous and not reported] | severe COVID-19 [Hospital admission up to 14 days after positive test] | Medical Research Council, UK Research and Innovation, Department of Health and Social Care, National Institute for Health Research and Community Jameel, Engineering and Physical Sciences Research Council | not declared |
| Sheikh, 2021 (preprint) (18) | United Kingdom | test-negative | 01.11.2021 - 19.12.2021 | general population, n=323648  [≥16 years of age, including previously infected] | Omicron (B.1.1.529), Delta (B.1.617.2) [sequencing/s-gene target failure] | any vaccine | full primary immunization, booster immunization [not reported] | symptomatic COVID-19 | RC, BREATHE—The Health Data Research Hub for Respiratory Health, UK Research and Innovation, Public Health Scotland, the Scottish Government Director General Health and Social Care and the University of Edinburgh | not declared |
| Spensley, 2022 (preprint) (19) | United Kingdom | test-negative | 01.12.2021 - 16.01.2022 | hemodialysis patients, n=1221  [median age: test-positive: 61 (IQR 49-74); test-negative: 66 (IQR 56-76), including previously infected] | Omicron (B.1.1.529) [sequencing/s-gene target failure] | Comirnaty, Vaxzevria | full primary immunization, booster immunization [homologous, other] | infection (any type), severe COVID-19 [hospitalization] | National Institute for Health Research Biomedical Research Centre, the Nan Diamond Fund, Sidharth and Indira Burman, the Auchi Charitable Foundation. | not declared |
| Tartof, 2022 (preprint) (20) | United States of America | test-negative | 01.12.2021 - 11.01.2022 | general population, n=14137  [adults, mean age: 52 (IQR 35-71), including previously infected] | Omicron (B.1.1.529), Delta (B.1.617.2) [dominance] | Comirnaty | full primary immunization, booster immunization [homologous] | severe COVID-19 [">=1 COVID-19 symptom and a positive KPSC laboratory-confirmed SARS-CoV-2 PCR test from a sample collected within 14 days prior to the initial admission date through 3 days after the admission"] | Pfizer Inc | yes |
| Thompson, 2022  (21) | United States of America | test-negative | 26.08.2021 - 05.01.2022 | general population, n=222772 (204745 during Delta dominance, 18027 during Omicron dominance)  [adults] | Omicron (B.1.1.529), Delta (B.1.617.2) [dominance] | Comirnaty, Spikevax | full primary immunization, booster immunization [not reported] | severe COVID-19 [laboratory-confirmed COVID-19–associated emergency department and urgent care encounters and hospitalizations] | CDC | yes |
| Tseng, 2022 (preprint) (22) | United States of America | case-control | 12.06.2021 - 31.12.2021 | general population, n=136345  [adults, mean age: test positive: 42,31 (SD ±14,64); test negative: 42,60 (SD ±14,67), including previously infected] | Omicron (B.1.1.529), Delta (B.1.617.2) [sequencing/s-gene target failure] | Spikevax | full primary immunization, booster immunization [homologous] | infection (any type), severe COVID-19 ["hospitalisation with SARS-CoV-2 positive test or hospitalization ≤7 days after a SARS-CoV-2 positive  test. COVID-19 hospitalization was confirmed by manual chart review conducted by a  physician investigator [BKA] to verify the presence of severe COVID-19 symptoms."] | Moderna Inc | yes |
| UK Health Security Agency, 2021; Andrews, 2021 (23, 24) | United Kingdom | test-negative | 27.11.2021 - 19.01.2022 | general population, n= not reported (fully primary vaccinated n=1042 316; booster-vaccinated n= 1288031; non-vaccinated n=229239)  [adults, including previously infected] | Omicron (B.1.1.529), Delta (B.1.617.2) [sequencing/s-gene target failure] | Comirnaty, Spikevax, Vaxzevria | full primary immunization, booster immunization [homologous and heterologous] | symptomatic COVID-19, severe COVID-19  [hospitalization] | no funding received | not declared |
| Veneti, 2022 (25) | Norway | cohort study | 06.12.2021 - 20.01.2022 | general population, n=91005 (51481 Delta cases, 39524 Omicron cases) | Omicron (B.1.1.529), Delta (B.1.617.2) [sequencing/s-gene target failure] | any vaccine | full primary immunization, booster immunization [not reported] | severe COVID-19 [hospitalisation ≤ 2 days before and ≤ 28 days following a positive COVID-19 test, where COVID-19 was the reported main cause of admission] | no funding received | not declared |
| Willett, 2022 (preprint) (26) | United Kingdom | test-negative | 22.12.2021 - 28.12.2021 | general population, n=101310  [adults, median age: test-positive: 44,73 (IQR 29,75-61,13); test-negative: 57,88 (IQR 40,54-68,7), including previously infected] | Omicron (B.1.1.529), Delta (B.1.617.2) [sequencing/s-gene target failure] | Comirnaty, Spikevax, Vaxzevria | full primary immunization [homologous], booster immunization [other] | infection (any type) | HDR-UK (E.C.T). COG-UK, the National Institute of Health Research, Genome Research Limited, Medical Research Council, G2P-UK National Virology Consortium | not declared |
| Young-Xu, 2022 (preprint) (27) | United States of America | test-negative | 01.12.2021 - 14.01.2022 | veterans, n=69215 (33% fully primary vaccinated, 31% booster vaccinated, 30% non-vaccinated) | Omicron (B.1.1.529), Delta (B.1.617.2) [dominance] | any vaccine | full primary immunization, booster immunization [not reported] | infection (any type) | US Food and Drug Administration, U.S. Department of Veterans Affairs Office of Rural Health. | not declared |

### Part 5: Effectiveness of COVID-19 vaccines against SARS-CoV-2 Delta variant infection (infection (any type), symptomatic COVID-19, and severe COVID-19 (hospitalization, ICU admission, or death))

|  | **Vaccine(s) used for immunization** | **Range of adjusted vaccine effectiveness (95% CI)^a,b,c^** | | | |
| --- | --- | --- | --- | --- | --- |
|  |  | **≈ 14 days^d^** | **>14 days up to 3 months** | **>3 months up to 6 months** | **>6 months** |
| **Infection (any type)** | | | | | |
| **Full primary immunization** | mRNA-based (any)^e^ | 41% (37–44) to 88.2% (83.1–91.8);  (5, 10, 11, 22, 26, 27) | 72.2% (70.4–74) to 87.8% (NR–NR);  (10, 22, 26) | 53.8% (52.9–54.6) to 80% (79–81);  (5, 10, 22) | 57.5% (50.4–63.3) to 71% (66–75);  (5, 22) |
|  | Vector-based (any)^f^ | 78.92% (NR–NR); (26) | 78.9% (NR–NR);  (26) | .. | .. |
|  | Heterologous scheme^g^ | .. | .. | .. | .. |
|  | Any vaccine^h^ | 31% (27–34) to 95% (94–95);  (3, 12, 15) | .. | .. |  |
| **Booster immunization** | mRNA-based (any)^e^ | 62% (38–78) to 93% (90–96)^i^;  (5, 10, 11, 22, 27) | 81.2% (79.2–82.9) to 82.8% (58.8–92.9)^i^; (10) | .. | .. |
|  | Vector-based (any)^f^ | .. | .. | .. | .. |
|  | Heterologous scheme^j^ | .. | .. | .. | .. |
|  | Any vaccine^h^ | 85.94% (NR–NR) to 97% (94–98);  (3, 26) | .. | .. | .. |
| **Symptomatic COVID-19** | | | | | |
| **Full primary immunization** | mRNA-based (any)^e^ | 90.0% (89.6–92.0) to 94.5% (90.5–96.9);  (23) | 85.5% (84.5–86.5) to 91.8% (89.6–93.6); (23) | 67.4% (66.5–68.2) to 76.2 (74.7–77.7);  (23) | 62.7% (61.6–63.7) to 80.4% (67.3–88.2);  (23) |
|  | Vector-based (any)^f^ | 82.8% (74.5–88.4);  (23) | 76.5% (70.3–81.5%); (23) | 47.4% (46.2–48.5);  (23) | 43.5% (42.4–44.5); (23) |
|  | Heterologous scheme^g^ | .. | .. | .. | .. |
|  | Any vaccine^h^ | 89% (86 – 92); (5)^k^ | 89% (87-90); (5)^k^ | 88% (87-89); (5)^k^ | 80% (74-84); (5) |
| **Booster immunization** | mRNA-based (any)^e^ | 83% (81–84) to 97% (96-98)^i^;  (1, 2, 5, 23) | 91.8% (91.4–92.1) to 96.4% (91.4–98.5); (23) | .. | .. |
|  | Vector-based (any)^f^ | .. | .. | .. | .. |
|  | Heterologous scheme^j^ | 95.4% (95.1–95.6) to 97.0% (96.7–97.3);  (23) | 92.6% (92.2–92.9) to 94.9% (93.8–95.9); (23) | .. | .. |
|  | Any vaccine^h^ | 77% (74–80) to 97% (96-98) ^i^;  (5, 18) | .. | .. | .. |
| **Severe COVID-19 (hospitalization, ICU admission, or death)** | | | | | |
| **Full primary immunization** | mRNA-based (any)^e^ | 76% (69–81) to 98.5% (92–99.7);  (7, 13, 14, 17, 21-23, 28) | 75% (63–84) to 99.2% (94.5–99.9); (17, 23) | 77% (62–86) to  98.3% (93.1–99.6);  (17, 23, 28) | 74% (65–80) to  95.3% (93.9–96.5);  (17, 21, 23, 28) |
|  | Vector-based (any)^f^ | 69% (-147–96) to 82% (50–93);  (14, 17) | 39% (-31–72) to  63% (-187–95);  (17) | 77% (22–93) to  92.9% (91.3–94.2);  (17, 23) | 90.6% (89.3–91.8); (23) |
|  | Heterologous scheme^g^ | 8% (-189–71); (14) | .. | .. | .. |
|  | Any vaccine^h^ | 47% (36–56) to 94% (84-98);  (5, 8, 12, 25) | 98% (94-99); (5) | 99% (98-99); (5) | 84% (79–88) to 95% (85-99);  (5, 25) |
| **Booster immunization** | mRNA-based (any)^e^ | 79% (58–90) to 99.6% (95.7–100);  (5, 13, 14, 21-23, 28, 29) | 84% (77–89);  (17) | .. | .. |
|  | Vector-based (any)^f^ | 87% (82–90) to 87% (84–88);  (17) | 88% (85–91);  (17) | .. | .. |
|  | Heterologous scheme^j^ | 99.4% (99.1–99.6) to 99.6% (98.9–99.8);  (23) | .. | .. | .. |
|  | Any vaccine^h^ | 88% (83–91) to 99% (98-99);  (5, 25) | .. | .. | .. |

**Footnotes:**

^a^ If not indicated otherwise, vaccine effectiveness (VE) estimates refer to the comparison of vaccinated (2 or 3 doses, respectively) vs. unvaccinated;

^b^ Provided VE estimates refer to the last reported timepoint per observation period (e.g. if studies reported VE after 2-4 and 5-9 weeks, the estimate of 5-9 weeks was included in the depicted effect range;

^c^ Several effect ranges are derived from single studies providing data for different vaccine types

^d^ timepoint closest to 14 days

^e^ Comirnaty or Spikevax

^f^ Vaxzevria or Janssen

^g^ 1^st^ dose with vector-based vaccine followed by prime booster dose of mRNA vaccine

^h^ Studies include recipients of different vaccine types, and data was not further stratified

^i^ Includes at least one study comparing booster vs. primary vaccination schedules (i.e., 3 vs. 2 doses)

^j^ 1^st^ dose with vector-based vaccine followed by prime booster dose, and 3^rd^ dose of mRNA vaccine OR vector-based primary vaccination followed by one booster dose of mRNA

^k^ One study estimated VE against vaccinated individuals who received second dose before ≥25 weeks, because of insufficient unvaccinated individuals available for analysis (18). VE against symptomatic infection with Omicron and Delta was estimated for 16-49 and 50+ year old’s, respectively. Estimates are provided for all reported observation periods in supplementary table 1

### Part 6: Characteristics of studies included for data extraction on VE against infection with the SARS-CoV-2 Omicron variant compared to infection with the SARS-CoV-2 Delta variant

| Author, year of publication | Study location | Study design | Study period | Study population, n [characteristics] | Variant analysed [method used to identify variant] | Vaccine analysed | Vaccine schedule analysed | Outcome analysed [definition of severe COVID-19 applied] | Funding received | Conflict of interest declared |
| --- | --- | --- | --- | --- | --- | --- | --- | --- | --- | --- |
| Davies, 2022 (preprint)  (8) | South Africa | cohort study | 14.11.2021 - 11.12.2021 | general population, n=9499 (5144 during Omicron period; 4355 during Delta period)  [64% 20-39 year olds; 17% 40-49 year olds; 11% 50-59 year olds; 5% 60-69 year olds, 2,7% >70 year olds, including previously infected] | Omicron (B.1.1.529) compared to Delta (B.1.617.2) [dominance] | Comirnaty, Janssen | full primary immunization [not reported] | severe COVID-19 [“Hospitalization included admission within 14 days before or after a COVID-19 diagnosis except for admissions to long term psychiatric or rehabilitation facilities”] | Western Cape Provincial Health Data Centre from the Western Cape Department of Health, the US National Institutes for Health, the Bill and Melinda Gates Foundation, the United States Agency for International Development, the European Union, the Wellcome Trust, the Medical Research Council of South Africa | none declared |
| Eggink, 2022  (30) | Netherlands | case-only study | 22.11.2021 - 19.01.2022 | general population, n= 174349 tests administered  [≥12 year olds, not previously infected] | Omicron (B.1.1.529) compared to Delta (B.1.617.2) [sequencing/s-gene target failure] | any vaccine | full primary immunzation | infection (any type) | not reported | not declared |
| Ferguson, 2021  (31) | United Kingdom | case-case study | 23.11.2021 - 11.12.2021 | general population, n= not reported (control group n=51263) [all agegroups, not previously infected] | Omicron (B.1.1.529) compared to Delta (B.1.617.2) [sequencing/s-gene target failure] | Comirnaty, Vaxzevria | full primary immunization, booster immunization | symptomatic COVID-19 | UK MRC and UKRI for Centre and research grant funding, the UK NIHR for Health Protection Research Unit funding, and Community Jameel for Institute and research funding | not reported |
| Hussey, 2022 (preprint)  (32) | South Africa | cohort study | 01.11.2021 - 14.12.2021 | general population, n=1636 [≥15 year olds, median 33 (IQR 25-44 "in those with RTD"; IQR 26-44 "in those without"), including both previously infected and non-infected] | Omicron (B.1.1.529) compared to Delta (B.1.617.2) [sequencing/s-gene target failure] | any vaccine | full primary immunization | severe COVID-19  [„individual tested positive for SARS-CoV-2 14 days before admission, or 7 days after the date of  admission, and was not admitted to a specialized psychiatric and rehabilitation facilities”] | not reported | not reported |
| Kahn, 2022 (preprint)  (12) | Sweden | test-negative | 27.12.2021 - 09.01.2022 | general population, n=55269  [including previously infected] | Omicron (B.1.1.529), Delta (B.1.617.2) [dominance] | any vaccine | full primary immunization, booster immunization [not reported] | infection (any type), severe COVID-19  [“at least 24 hour-hospitalization five days before until 14 days after a positive test and with a need of oxygen supply ≥ 5 L/min or admittance to an intensive care unit”] | Swedish Research Council, Sweden’s Innovation Agency,  internal grants for thematic collaboration initiatives at Lund University | not declared |
| Kislaya, 2022 (preprint)  (33) | Portugal | case-case study | 06.12.2021 - 26.12.2021 | general population, n=13143 (4898 during Omicron period, 8245 during Delta period) [≥12 year olds, not previously infected] | Omicron (B.1.1.529) compared to Delta (B.1.617.2) [sequencing/s-gene target failure] | any vaccine, Comirnaty, Vaxzevria | full primary immunization, booster immunization | infection (any type) | no funding received | none |
| Peralta Santos, 2022 (preprint)  (34) | Portugal | cohort study | 01.12.2021 - 29.12.2021 | general population, n= 15978 (6581 Omicron cases, 9397 Delta cases) [≥16 years of age, mean 37,1 (SD 14,8) (Omicron period), 43,4 (SD 15,8) (Delta period), including both previously infected and non-infected] | Omicron (B.1.1.529) compared to Delta (B.1.617.2) [sequencing/s-gene target failure] | any vaccine | full primary immunization, booster immunization | severe COVID-19  [„any record of death on the national Death Certificate Information System with COVID-19 as the primary cause of death (ICD-10 code U.071) according to the WHO classification”, “admission to a public hospital in Portugal mainland within the 14 days following a positive sample SARS-CoV-2 collection, notified in the surveillance system”] | not reported | not reported |
| Veneti, 2022  (25) | Norway | cohort study | 06.12.2021 - 20.01.2022 | general population, n=91005 (51481 Delta cases, 39524 Omicron cases) | Omicron (B.1.1.529), Delta (B.1.617.2) [sequencing/s-gene target failure] | any vaccine | full primary immunization, booster immunization [not reported] | severe COVID-19  [hospitalisation following a positive SARS-CoV-2 test] | no funding received | not declared |

### Part 7: Effect measures of vaccine effectiveness against infection with the SARS-CoV-2 Omicron variant compared to infection with the SARS-CoV-2 Delta variant for the outcomes infection (any type), symptomatic COVID-19, and severe COVID-19 (hospitalization, ICU admission, or death)

| Author, year of publication | Intervention | Vaccine | Time point of analysis after vaccination | Outcome  [definition of severe COVID-19 applied] | Effect measure | # cases Delta, among intervention group | # cases Omicron, among intervention group | Point estimate | CI, low | CI, high |
| --- | --- | --- | --- | --- | --- | --- | --- | --- | --- | --- |
| Davies, 2022  (8) | full primary immunization | any vaccine (Janssen/ Comirnaty) | >28 days after Janssen vaccine, >14 days after Comirnaty vacccine | severe COVID-19 [„Deaths within 14 days of COVID-19 diagnosis were included unless a clear non-COVID-19 cause of death was recorded"] | HR | not reported  (4355 total cases) | not reported (5104 total cases) | 0.20 | 0.09 | 0.43 |
|  |  |  |  | severe COVID-19 ["severe hospitalisation (=Admission to an intensive care unit, mechanical ventilation or prescription of oral or intravenous steroids;) and death"] | HR | not reported (4355 total cases) | not reported (5104 total cases) | 0.23 | 0.13 | 0.39 |
|  |  |  |  | severe COVID-19 ["hospitalization/death"] | HR | not reported (4355 total cases) | not reported (5104 total cases) | 0.42 | 0.34 | 0.52 |
| Eggink, 2022  (30) | full primary immunization | any vaccine | >14 days after 2nd vaccine/>28 days after 1 dose of Janssen vaccine | infection (any type), overall | OR | not reported (57674 cases among fully primary immunized) | not reported (53291 cases among fully primary immunized) | 3.6 | 3.4 | 3.7 |
|  |  |  |  | infection (any type), in people without previous travel history | OR | not reported (1000 cases among people without previous travel history) | not reported (4162 cases among people without previous travel history) | 3.4 | 3.2 | 3.5 |
|  |  |  |  | infection (any type), in 12-29 year olds | OR | not reported (25245 cases among 12-29 year olds) | not reported (43907 cases among 12-29 year olds) | 4.1 | 3.9 | 4.4 |
|  |  |  |  | infection (any type), in 30-59 year olds | OR | not reported (48771 cases among 30-59 year olds) | not reported (32592 cases among 30-59 year olds) | 3.2 | 3 | 3.4 |
|  |  |  |  | infection (any type), in ≥ 60 year olds | OR | not reported (19718 cases among ≥ 60 year olds) | not reported (4116 cases among ≥ 60 year olds) | 2.8 | 2.3 | 3.2 |
| Ferguson, 2021  (31) | full primary immunization | Vaxzevria | >14 days | symptomatic COVID-19 | HR | 32887 | 1676 | 1.86* | 1.74 | 1.98 |
|  | booster immunization |  | <14 days |  | HR | 4926 | 250 | 1.86* | 1.67 | 2.08 |
|  | booster immunization |  | >14 days |  | HR | 1192 | 230 | 4.32* | 3.84 | 4.85 |
|  | full primary immunization | Comirnaty | >14 days |  | HR | 17544 | 2888 | 2.68* | 2.54 | 2.83 |
|  | booster immunization |  | <14 days |  | HR | 890 | 60 | 2.49* | 2.06 | 3.01 |
|  | booster immunization |  | >14 days |  | HR | 1801 | 288 | 4.07* | 3.66 | 4.51 |
| Hussey, 2022  (32) | full primary immunization | any vaccine (Janssen/ Comirnaty) | ≥28 days after Janssen/≥14 days after Comirnaty | severe COVID-19 [„individual tested positive for SARS-CoV-2 14 days before admission, or 7 days after the date of admission, and was not admitted to a specialized psychiatric and rehabilitation facilities, it was defined as COVID-19 related hospital admission”] | HR | 33 (150 total cases) | 312 (1486 total cases) | 0.45 | 0.26 | 0.77 |
| Kahn, 2022  (12) | full primary immunization | any vaccine | not reported | severe COVID-19 [„at least 24 hour-hospitalization five days before until 14 days after a positive test and with a need of oxygen supply ≥ 5 L/min or admittance to an intensive care unit (ICU)”] | OR | 4031 | 21678 | 0.29* | 0.18 | 0.46 |
| Kislaya, 2022  (33) | full primary immunization | any vaccine | >14 days | infection (any type) | OR | 7245 | 4515 | 2.1 | 1.8 | 2.4 |
|  |  |  | <113 days |  | OR | 1141 | 1002 | 2.3 | 1.9 | 2.8 |
|  |  |  | 113-168 days |  | OR | 4313 | 2640 | 2.0 | 1.7 | 2.4 |
|  |  |  | ≥169 days |  | OR | 1791 | 873 | 1.9 | 1.6 | 2.3 |
|  |  | Comirnaty | >14 days | infection (any type) | VE | not reported | not reported | 28.1* | 12.2 | 40.9 |
|  |  | Vaxzevria | >14 days |  | VE | not reported | not reported | -7.1* | -31 | 11.9 |
|  |  | any vaccine | >14 days | infection (all), ≥ 50 year olds | OR | 2089 | 746 | 1.7 | 1.1 | 2.7 |
|  | booster immunization | any vaccine | <14 days | infection (all), ≥ 50 year olds | OR | 283 | 97 | 2.1 | 1.3 | 3.6 |
|  |  |  | >14 days |  | OR | 159 | 149 | 5.2 | 3.1 | 8.8 |
|  |  | Comirnaty | >14 days |  | VE | not reported | not reported | 68.8* | 46.4 | 81.7 |
|  |  | Vaxzevria | >14 days |  | VE | not reported | not reported | 64.2* | 36.8 | 79.4 |
| Peralta-Santos, 2022  (34) | full primary immunization | any vaccine | ≥14 days following 2^nd^ vaccine dose or 1^st^ vaccine dose of Ad26.COV2.S or up to 14 days after the booster dose uptake | Death [„any record of death on the national Death Certificate Information System (SICO) with COVID-19 as the primary cause of death (ICD-10 code U.071) according to the WHO classification”] | HR | not reported | not reported | 0.335 | 0.131 | 0.925 |
|  | full primary immunization + booster immunization |  |  |  | HR | not reported | not reported | 0.046 | 0.008 | 0.195 |
|  | full primary immunization + booster immunization |  |  | severe COVID-19 [„admission to a public hospital in Portugal mainland within the 14 days following a positive sample SARS-CoV-2 collection, notified in the surveillance system”] | HR | not reported (148 total cases) | not reported (16 total cases) | 0.34 | 0.13 | 0.87 |
| Veneti, 2022  (25) | full primary immunization | any vaccine | 7–179 days after second dose | hospitalisation [„hospitalisation following a positive SARS-CoV-2 test”] | HR | 47 | 26 | 0.62 | 0.31 | 1.24 |
|  |  |  | ≥ 180 days after second dose |  | HR | 98 | 26 | 0.5 | 0.25 | 1.01 |
|  |  |  | ≥ 7 days after third dose |  | HR | 55 | 20 | 0.19 | 0.08 | 0.43 |

*unadjusted effect estimate
